# Proteomic signatures of pediatric cardiovascular and cardiometabolic traits demonstrate long-term modifiable drivers of adult disease

**DOI:** 10.1101/2025.10.27.25338754

**Authors:** Joshua M Landman, Heather M Highland, Andrew S Perry, Annie G Howard, Quanhu Sheng, Anna Lorenz, Alexandra B Palmer, Shilin Zhao, Wanying Zhu, Xinruo Zhang, Victoria L Buchanan, Elizabeth G Frankel, Rashedeh Roshani, Alyssa Scartozzi, Eric H Farber-Eger, Mohammad Y Anwar, Jessica K Sprinkles, Ash Breidenbach, Ting-Chen Wang, Christine A Ballard, Matthew Nayor, Jaclyn Tamaroff, Absalon Gutierrez, Lauren E Petty, Alexander S Petty, Benjamin Lippi, Lindsay Fernandez-Rhodes, Hung-Hsin Chen, Mohanraj Krishnan, Mariaelisa Graff, Katie A Meyer, Miryoung Lee, Kristin L Young, Quinn Wells, Jane E Freedman, Eric R Gamazon, Joseph B McCormick, Susan P Fisher-Hoch, Penny Gordon-Larsen, Jennifer E Below, Kari E North, Ravi V Shah

## Abstract

While the earliest pathological signs of cardiovascular disease (CVD) emerge before age 20, current adult-based risk thresholds fail to identify a substantial fraction of high-risk children. With the rising prevalence of childhood obesity, the need for early and sensitive detection of cardiovascular-kidney-metabolic disease (CKMD) risk is paramount to enable timely intervention that can prevent the trajectory toward CVD in later life. To identify early accessible molecular biomarkers of CKMD in children, we measured 25 CKMD phenotypes, spanning liver, adipose, vascular, and dysglycemia traits, linked those to the circulating proteome and built a multi-protein signature of composite CKMD, leveraging data from 273 children and adolescents (13.1 ± 2.7 years; 53% females). We compared these results to the adult CKMD proteome, using data from 685 adults from the same community and 28,257 adults from the UK Biobank. The pediatric CKMD proteome was highly concordant with the adult CKMD proteome, reflecting known and novel mechanisms of pancreatic beta-cell health and insulin sensitivity, liver homeostasis, inflammation, and cholesterol metabolism. Similarly, multi-protein signatures of composite CKMD phenotypes in children were strongly associated with CKMD-related outcomes in both adult populations. Importantly, many proteins linked to pediatric CKMD were modifiable with GLP-1 receptor agonist therapy, associated with adult CKMD-related outcomes in a large-scale proteome-wide association study (PWAS), and exhibited significant variability during development (ages 4-24 years). These findings demonstrate that CKMD develops starting early in life-course continuum, with proteomic profiles linked to future irreversible CVD conditions emerging early in life when they may still be reversible. This highlights a critical opportunity to improve CVD-free longevity through precision medicine diagnosis and intervention in children and adolescents.

## INTRODUCTION

The earliest pathological signs of cardiovascular disease (CVD) may emerge before age 20,^1^ driven by cumulative exposure to obesity, insulin resistance, hypertension, and pro-atherogenic dyslipidemia.^2^ Despite strong epidemiologic evidence highlighting the importance of these cardiovascular and metabolic risks in premature CVD,^3–8^ current CVD prevention efforts remain adult-centered, missing an opportunity to identify high-risk pediatric individuals. Most children and adolescents harbor intermediate, subclinical phenotypes that fall below adult thresholds. Nevertheless, even “normal” values in adolescents are linked to later CVD in adulthood.^3,4^ For example, screening tests commonly used in adults (e.g., fasting glucose) miss a substantial fraction (up to 70%)^9^ of dysglycemia and type 2 diabetes in children^10^. In adults, molecular biomarkers have been advanced as a “precision” approach to detect disease liability decades before clinical onset,^11–14^ yet analogous studies in pediatric populations are lacking.

Linking pediatric biomarkers to future clinical risk requires longitudinal follow-up data, further challenging efforts to identify reversible, actionable, prognostic childhood risk mechanisms.

In this study, we linked 25 cardiovascular-kidney-metabolic disease (CKMD) phenotype in 273 Hispanic/Latino children (mean age 13.1 ± 2.7 years; 53% female) to the circulating proteome to identify molecular biomarkers of early CVD susceptibility in children. CKMD phenotypes spanned clinical and imaging-based measures known to influence CVD risk in adults, including body composition, liver fat and fibrosis, renal function, and traditional CVD risk factors. We identified proteomic patterns of these phenotypes in children at single- and multi-trait levels, comprising both putative CVD pathways and composite multi-CKMD phenotype proteomic signatures that capture shared molecular risk, reflecting their high interrelatedness, frequent co-occurrence, and many notable shared mechanistic pathways. We subsequently mapped the pediatric multi-protein composite CKMD signature to adult CVD in two samples: (1) 494 Hispanic/Latino adults from the same community (mean age 51 ± 14.3 years; 68% female) and (2) 28,257 adults from the UK Biobank (mean age 58 ± 8 years; 54% female). To assess modifiability, we next evaluated whether proteins implicated in pediatric CKMD can be modified by glucagon-like peptide-1 receptor agonist (GLP-1RA) therapy in the STEP 1 randomized clinical trial.^15^ In parallel, we constructed genetic instruments^16^ of the circulating proteome to conduct proteome-wide association studies (PWAS) to test whether this GLP1-RA-modifiable pediatric CKMD proteome would exhibit a life-course, potentially causal susceptibility to adult CKMD (paralleling studies in UK Biobank). Finally, we studied sources of variability in the pediatric CKMD-associated proteome during development across the first two decades of life through recently published resources^17^, a prerequisite to intervention. Ultimately, our goal was to link pediatric CKMD phenotypes to long-term adult CVD through integrated deep phenotyping, proteomic, and genomic studies across the life-course, opening a window into the earliest roots of CVD at its most interruptible phase.

## METHODS

### Study sample

The Border Health Research Cohort - Cameron County (BHRC, formerly Cameron County Hispanic Cohort, CCHC) study is a longitudinal, population-based cohort study of individuals from randomly ascertained households in Cameron County, Texas, located on the US-Mexico border. Pediatric participants aged 8-17 were enrolled from participating BHRC households. Their study exams were included in the analysis if they were under 18 years at exam 1; when available, follow-up exams up to age 20 were included. Of the 607 BHRC youth exams potentially eligible for this analysis, 397 included whole blood proteomic profiling. Of these, 3 were excluded due to poor sample quality, leaving a final analytic sample of 394 (273 individuals at exam 1 and 121 individuals at exam 2). Analyses for individual phenotypes include fewer exams for some traits based on data completeness (see **Table 1** for total N per trait).

**Table 1:** Demographic summary of BHRC children and adults in proteomics datasets. Traits analyzed and clinical categorizations are listed as either mean (SD) or N (%).

| Domain | Trait (units) | Abbreviation | Multi-trait analysis | Pediatric Mean (SD) | Pediatric N<sub>ind</sub> | Adult Mean (SD) | Adult N<sub>ind</sub> |
| --- | --- | --- | --- | --- | --- | --- | --- |
|  | Age (years) |  | No | 13.1 (2.7) | 273 | 51.5 (14.3) | 494 |
|  | Sex, n (% female) |  | No | 145 (53) | 273 | 356 (68.3) | 494 |
| *Dysglycemia* | Fasting blood glucose (mg/dL) | FBG | Yes | 88.63 (6.97) | 270 | 92.2 (8.8) | 289 |
|  | HbA1c, % | HbA1c | Yes | 5.34 (0.29) | 271 | 5.54 (0.32) | 245 |
|  | HOMA-IR | HOMA-IR | Yes | 3.1 (1.9) | 260 | 2.4 (1.6) | 259 |
| *Blood pressure* | Diastolic blood pressure (mmHg) | DBP | No | 62.2 (7) | 273 | 73.8 (9.1) | 486 |
|  | Systolic blood pressure (mmHg) | SBP | Yes | 102.8 (10) | 273 | 122.9 (19.2) | 486 |
|  | Pulse pressure (mmHg) | PP | No | 40.7 (6.7) | 273 | 48.8 (14.4) | 486 |
|  | Mean arterial pressure (mmHg) | MAP | No | 75.7 (7.5) | 273 | 89.7 (11.5) | 486 |
| *Kidney* | eGFR (sex- and age-dependent equation) | eGFR | Yes | 108.7 (18.6) | 270 | 98.7 (18.7) | 465 |
| *Dyslipidemia* | High-density lipoprotein (mg/dL) | HDL-C | Yes | 48.4 (10.7) | 271 | 50.1 (14.5) | 491 |
|  | Low-density lipoprotein (mg/dL) | LDL-C | Yes | 78.2 (22.5) | 271 | 105.2 (32.5) | 484 |
|  | Total cholesterol (mg/dL) | TC | Yes | 146.0 (26.1) | 271 | 183.3 (37.4) | 491 |
|  | Triglycerides (mg/dL) | TG | Yes | 97.4 (52.5) | 271 | 145.3 (92.4) | 491 |
| *Liver* | Alanine Aminotransferase (U/L) | ALT | Yes | 26.0 (21.0) | 271 | 32.5 (18.3) | 473 |
|  | Aspartate Aminotransferase to Platelet Ratio Index | APRI | Yes | 0.24 (0.14) | 271 | 0.32 (0.22) | 464 |
|  | Aspartate Aminotransferase (U/L) | AST | Yes | 21.4 (10.7) | 271 | 24.4 (13.2) | 477 |
|  | Controlled Attenuation Parameter (dB/m) | CAP | No | 245.8 (57.8) | 148 | 288.1 (63.2) | 437 |
|  | Fibrosis-4 | FIB | Yes | 0.23 (0.1) | 271 | 0.97 (0.56) | 462 |
|  | Liver Stiffness Measurement (kPa) | kPa | No | 4.46 (1.57) | 162 | 5.96 (4.67) | 429 |
| *Adiposity* | BMI z-score | BMI-Z | Yes | 0.96 (1.18) | 273 | NA | NA |
|  | BMI (kg/m<sup>2</sup>) |  | No | NA | NA | 32.8 (8.9) | 494 |
|  | Total fat mass (g) | TFM | No | 21,563<br>(10,670) | 148 | 32161<br>(11391) | 356 |
|  | Total percent fat | TPF | No | 36.0 (9.3) | 148 | 41.21 (8.98) | 356 |
|  | Total lean mass excluding bone mass (g) | TLM | No | 34,712<br>(10,555) | 148 | 42942<br>(10577) | 356 |
|  | Total percent lean excluding bone mass | TPL | No | 60.8 (8.9) | 148 | 55.97 (8.66) | 356 |
|  | Trunk fat (g) | TrF | No | 9,475<br>(5,340) | 148 | 15964<br>(5768) | 356 |
|  | Visceral fat area (cm<sup>2</sup>) | VFA | No | 73 (38) | 148 | 199 (80) | 356 |
| *Clinical classifications* | Hypertension (SBP ≥ 130 mmHg, DBP ≥ 80 mmHg, or taking anti-hypertensive meds), % |  | No | 2 (0.70) | 273 | 232 (45.3) | 494 |
|  | Low HDL (< 50 mg/dL females, <40 mg/dL males), % |  | No | 105 (38.5) | 273 | 218 (44.4) | 494 |
|  | High triglyceride (> 150 mg/dL or on lipid lowering medication), % |  | No | 35 (12.8) | 273 | 248 (50.4) | 494 |
|  | Obese (BMI percentile > 95<sup>th</sup>% children, BMI > 30 kg/m<sup>2</sup> adults), % |  | No | 95 (34.8) | 273 | 269 (54.1) | 494 |
|  | Diabetes (diagnosed type 2 diabetes, on diabetes medication, FBG > 126 mg/dL, or HbA1c ≥ 6.5%), % |  | No | 1 (0.4) | 273 | 207 (39.8) | 494 |

Adults from the same community aged 18-98, were enrolled and followed longitudinally every 5 years. Of the 4,956 adult BHRC participants, 887 exams from 686 individuals had proteomics available and were included in analyses validating associations in adults; 494 participants were included for testing the fit of pediatric models to adult phenotypes. No participants included in the pediatric analyses were included in the internal adult replication.

Pediatric assents/consents and parental/guardian consents were obtained according to the approved IRB protocol (HSC-SPH-14-0141, UT Health Houston School of Public Health-Brownsville), including the use of optional bilingual instruments. Written informed consent was obtained from all participants under the approved IRB protocol.

### Pediatric and adult CKMD phenotyping

Descriptive statistics for each trait and outcome are reported in **Table 1**, with additional details available in **Supplemental Table 2.** Participants were examined in the Clinical Research Unit in Brownsville, Texas. Children were accompanied by a parent or guardian. Venipuncture was performed for fasting glucose, fasting insulin, glycated hemoglobin (HbA1c), lipid profile (high-density lipoprotein [HDL-C], total cholesterol [TC], and triglycerides [TG]), creatinine, and alanine and aspartate aminotransferase (ALT, AST). From these measures, we derived homeostatic model of insulin resistance (HOMA-IR),^18^ calculated low-density lipoprotein (LDL-C),^19^ TG/HDL ratio, estimated glomerular filtration rate (eGFR),^20^ AST-to-platelet ratio index (APRI), and fibrosis-4 index (FIB-4).^21^ Resting systolic and diastolic blood pressures (SBP, DBP) were measured three times, with the average of the last two measures used; mean arterial pressure (MAP) and pulse pressure (PP) were then calculated. Non-invasive ultrasound imaging of the liver using FibroScan (Echosens, Westborough, MA) was used to measure liver stiffness (kPA) and controlled attenuation parameter (CAP). Participants underwent whole-body dual energy X-ray absorptiometry (DXA) for body composition via standard clinical protocols (Hologic Inc., Marlborough, MA) to derive total and percent fat mass, total and percent lean mass (excluding bone mass), trunk fat, and visceral fat area (cm^2^). Weight and height were measured by study staff for BMI. Age and sex specific BMI-Z scores for children and adolescents were calculated based on the Centers for Disease Control and Prevention (CDC) growth charts^22^. Trait-specific QC strategies are described in **Supplemental Table 2.**

### Proteomics

We measured 5,420 circulating proteins (Olink Explore HT) in 394 frozen serum samples from children; one sample was removed due to poor quality. Normalized protein expression (NPX; protein abundance in log_2_ units) was calculated as indicated by the manufacturer (version 6.7.2 Olink proprietary software). Methods for Olink-based proteome quantification have previously been described.^23^ In adults, we used the same method and panel for 509 exams from 495 participants (“batch 2”), and an earlier panel (Olink Explore 3072 with 2,940 proteins) for 378 exams in 191 participants (“batch 1”). Overlapping proteins were mapped across the two batches for N-weighted meta-analysis. The subset of 494 participants in batch 2 was used for comparisons of effect size.

### Association of the circulating proteome with CKMD phenotypes in children and adults

We used linear mixed effects regression to estimate associations between proteins and CKMD phenotypes in children. HOMA-IR and triglyceride level were log-transformed to ensure normality. Each mixed effects regression model included a single phenotype as a function of single protein and included a participant random effect to account for repeated measurement of phenotypes over exams. In our primary analyses, we adjusted for age and sex. As a sensitivity analysis, we additionally adjusted for BMI-Z (or adult BMI), except for measures of body composition (e.g., DXA measures). Trait-specific modeling strategies are described in **Supplemental Table 2.** Type 1 error was controlled using the Benjaminini-Yekutieli false discovery rate (FDR) across all trait-protein comparisons.^24^ In adults, we applied the same modeling strategy (and additionally controlled for the first 3 genetic principal components) to measure the association between each phenotype and protein in adults (using standard deviation-scaled model coefficients). Effect sizes were compared to the pediatric results via Pearson correlation. Pathway analyses were performed using proteins associated with phenotypes (at 5% FDR in children), with all proteins used in regression analysis as the universe background on a KEGG^25^ and WikiPathways^26^ (FDR-adjusted P for enrichment was 0.1).

To assess curated evidence from published genome-wide association studies (GWAS), we classified phenotypes into 6 domains: dysglycemia (fasting glucose, HbA1c, HOMA-IR), body composition (BMI-Z, total fat mass (TFM), total percent fat (TPF), total lean mass excluding bone mass (TLM), total percent lean mass excluding bone mass (TPM), trunk fat mass (TrF), and visceral fat area (VFA)), blood pressure (SBP, DBP, MAP, PP), dyslipidemia (LDL-C, HDL-C, TC, TG, TG/HDL-C ratio), hepatic (ALT, AST, APRI, FIB-4, CAP, kPa), and renal (eGFR). We queried the top 30 proteins by standardized effect size in the pediatric analyses associated with each of the 6 phenotype domains at a 5% FDR (N=245 unique proteins), along with proteins associated with at least one individual phenotype in children but not in adults (N=109 proteins) in the Common Metabolic Diseases Knowledge Portal (CMDKP^27^). From the CMDKP, we used the Human Genetic Evidence (HuGE) scores for all phenotypes to summarize extant evidence of the corresponding gene’s association with 22 CMDKP cataloged phenotype groups. Evidence was grouped into bins of compelling (HuGE score≥350), extreme (≥100), very strong (≥30), strong (≥10), moderate (≥3), anecdotal (≥1), and no evidence (<1) for visualization in heatmaps^27^.

### Identifying a proteomic signature across CKMD phenotypes in childhood using machine learning approaches

We next investigated joint associations between the circulating proteome and 16 CKMD phenotypes across the 6 phenotype domains in 273 children with both proteomic and clinical data (see **Table 1**). Our selected approach required a complete, cross-sectional proteomic-phenotype dataset. Consequently, we excluded phenotypes analyzed in the single-trait analyses that had over 50% missingness across the union of all participants in those analyses, leaving us with only 11 individuals with any missing data. We imputed missing phenotype data for those 11 individuals via random forest imputation (*missForest*)^28^. Prior to imputation, TG and HOMA-IR were log-transformed, all hepatic traits were inverse rank normalized, and all proteins and phenotypes were mean-centered and standardized.

We performed principal components analysis (PCA) across the 16 phenotypes, selected six PCs by scree plot evaluation, and performed varimax post-rotation^29^. Using each of the six PCs as a dependent variable and our proteomic data as the independent variables, we used L1-regularized regression with repeated cross-validation (Least Absolute Shrinkage and Selection Operator, LASSO regression) to construct six parsimonious multivariable proteomic signatures. Model fit (R^2^) was evaluated by performance in cross-validation folds in *caret*^30^.

### Pediatric proteomic signatures of CKMD and long-term adult CKMD outcomes

We next measured the relationship between the LASSO-derived proteomic scores for children, corresponding to each of the six PCs, and adult cross-sectional and longitudinal phenotype outcomes. We estimated the proteomic scores for each multi-phenotype PC in adults from two samples: (1) BHRC adults (n=494) and (2) adults from UK Biobank (n=28,257)^31^.

Given the differences in coverage across Olink platforms, (Explore 3072 vs. HT) between pediatric and adult samples (from both the BHRC and the UK Biobank, which used HT and Explore 3072, respectively), we recalibrated our LASSO pediatric score for application to these datasets, following published methods^32^. In brief, we refit each of our six models for the subset of proteins available on the Explore 3072 platform using the pediatric proteomic scores for each phenotypic PC as the dependent variable and the subset of pediatric proteins common across samples as the independent variables. We confirmed adequate performance of the recalibrated LASSO model within the sample of children, with R^2^ values across PCs ranging from 0.72-0.98. Coefficients from the recalibrated LASSO-based PC proteomic models were applied to mean-centered and standardized protein abundance in the adult sample to generate adult PC proteomic scores for association analyses. PC proteomic scores were defined the linear combination of the pediatric LASSO coefficient and the standardized protein values across all LASSO-included proteins. Standardized scores were then used as an independent variable in Cox regression models for all-cause mortality and selected CKMD outcomes.

Our analytic sample in the UK Biobank included 28,257 unique participants, from which we parsed out six different subsets of participants (with participants allowed to be in >1 subset) based on having complete data for each set of proteins in each LASSO-based PC signature of pediatric CKMD. Clinical outcomes in the UK Biobank were defined using ICD codes summarized by the PheWAS package^33^. We excluded participants from models if a confounding ICD code was present or if they reported a prevalent diagnosis or related condition (UK Biobank Data Fields 2443, 20002). Time-to-death was defined by registry data (UK Biobank Data Field 40000), and non-deaths were censored on 30 November 2022. Time-to-event data for non-death outcomes was defined as the time to first qualifying ICD code, and non-event participants were censored at the date of death (if applicable) or region-specific dates (UK Biobank Data Field 54): 31 October 2022 for England; 31 July 2021 for Scotland; and 28 February 2018 for Wales. Cox regression models included adjustments to capture metabolic risk measures used in clinical practice and more general confounders for mortality: age, sex, race/ethnicity, BMI, HbA1c, TG, HDL-C, Townsend deprivation index, smoking, and alcohol use. We included non-linear terms as appropriate, given potential non-linear dependence of CKMD risk on age. Our methods were adapted with minimal modification from prior work in our group to ensure scientific reproducibility^34^.

### Dynamic changes in the circulating proteome with GLP-1 receptor agonist therapy

Proteins associated with pediatric CKMD that are dynamic under GLP-1RA therapy may identify actionable biomarkers of long-term risk and support utilization of GLP-1RA in at-risk children^35^. Therefore, we mapped 1,032 proteins related to any CKMD phenotype in our pediatric sample (in age- and sex-adjusted models at a 5% FDR) to a recently reported study of circulating proteins in the STEP 1 randomized clinical trial (semaglutide)^15^. We examined changes in the circulating proteome after 68 weeks of semaglutide therapy in the STEP 1 cohort (SomaScan aptamer-based platform), matching proteins identified in our cohort by UniProt identifier and restricting to those proteins with significant on-therapy change in STEP 1 (q-value ≤ 0.05). For duplicate aptamers for a given protein, the minimal absolute value effect size was chosen.

### Genetically predicted proteome-wide association analyses

To investigate the impact of modifiable CKMD proteins on adult cardiovascular disease (CVD) outcomes, we leveraged genetic prediction models using proteogenomic data from the Atherosclerosis Risk in Communities (ARIC) study (N=7,213, Somalogic aptamer-based proteomics)^36^. We applied proteome-wide association studies (PWAS)^16^ to identify proteins linked to adult CKMD outcomes, focusing on those previously associated with pediatric CKMD phenotypes and responsive to GLP-1RA treatment (FDR < 5%). We constructed protein expression models which were integrated into the Mendelian randomization (MR) joint-tissue imputation (JTI) pipeline to enable causal inference^37^.

Among the available 1,318 protein models (trained using elastic net), 131 proteins were significantly associated with pediatric CKMD phenotypes, and showed evidence of change after GLP-1RA, based on STEP 1 trial findings. To test whether genetically predicted protein levels of these 131 proteins are associated with CKMD clinical outcomes in adults, we leveraged published large-scale GWAS of heart failure^38^ (N=1,946,349 individuals from 42 studies, 153,174 cases), BMI (N=419,163 in UK Biobank^39^), type 2 diabetes^40^ (N=2,535,601 individuals, 428,452 cases), and renal function^41^ (creatinine; N=1,004,040). We estimated the effect (z-score) of the genetically predicted protein levels 𝑔 on each clinical outcome as follows^42,43^:

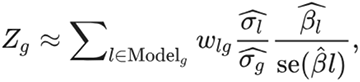

Here, 𝑤_i,g"_ is the weight assigned to the variant 𝑙 by the trained protein model, 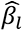 and se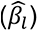 are the GWAS effect size and corresponding standard error summary statistics, respectively, and 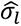 and 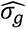 are the estimated variance for the variant 𝑙 and protein 𝑔, respectively.

### Sources of variability in early development

To investigate sources of variability in early development, we utilized very recent published data from a population-based birth cohort from Sweden (BAMSE)^17^. We extracted reported variance explained by age/sex and subject-specific random effect from linear mixed models analyzing Olink Explore-derived NPX protein levels in 50 males and 50 females measured longitudinally at ages 4, 8, 16, and 24^17^. We plotted variance explained across variance components for proteins associated with both pediatric and adult CKMD in our BHRC sample. We highlighted a selection of participant-level proteins across time in BAMSE from the Human Protein Atlas (https://www.proteinatlas.org).

## RESULTS

### Characteristics of pediatric and adult samples

Pediatric participants were predominantly in mid-adolescence (13.7 ± 2.8 years of age), with approximately 53% identifying as female (**Table 1**). More than a third of BHRC pediatric participants had obesity with indications of pro-atherogenic dyslipidemia, hypertension, and insulin resistance. Adult participants from the same community were at similarly high risk for these conditions (51.5 ± 14.3 years of age, 54.1% obese, 39.8% T2D, 68.3% females). We observed expected associations among CKMD phenotypes in children and adults, with body composition associated with greater insulin resistance, body fat distribution, pro-atherogenic dyslipidemia (high TG and low HDL-C), and elevated blood pressure (**Supplemental Figure 1**).

### Characterizing the pediatric CKMD proteome

Among proteins associated with glycemic control, we observed markers indexing distinct facets of β-cell function and insulin sensitivity. For example, C-peptide^10^ (an indicator of residual β-cell secretory reserve) emerged in our phenome associations, reinforcing the value of measuring endogenous insulin secretion even in early disease. Meanwhile, chromogranin B (CHGB), known to modulate insulin granule trafficking in β-cells by facilitating proinsulin transit from the trans-Golgi to the plasma membrane, was inversely related to dysglycemia in our pediatric cohort, echoing adult findings of CHGB influence on metabolic traits.^44–46^ Additionally, canonical adult markers of insulin sensitivity, such as GUSB^47^,IGFBP-1, and IGFBP-2, were detected and showed consistent directionality in children.^48,49^

For hepatic traits, significant proteins implicate core metabolic machinery in the liver. For example, GPD1 (glycerol-3-phosphate dehydrogenase) plays a central role in glycerolipid metabolism and gluconeogenesis,^50,51^ while ALDH1A1 (aldehyde dehydrogenase) is a key node in retinoid and aldehyde detoxification pathways.^52^ Some proteins, such as IGSF9, have been previously linked in adult cohorts to steatosis or fibrosis; ACY1/ACY2 likewise have been nominated in previous metabolic disease or hepatic injury studies.^49,53,54^ The detection of such proteins in children suggests either early onset of perturbations or latent predisposition to hepatic risk.

Our results span both “protective” and “risk” associations with the CKMD phenome, and many associations appear modulated by body mass.^55,56^ For instance, elevated creatine kinase B (CKB)—a key enzyme in phosphagen energy shuttling and implicated in thermogenesis in highly metabolic tissues—was associated with healthier adiposity phenotypes in children. This is consistent with murine knockout studies where CKB influences energy expenditure, inflammation in adipose tissue, and obesity outcomes.^57,58^ Similarly, CHGB’s association with lower dysglycemia fits with its secretory role and with adult metabolic directionality.^45^

We also confirmed prior findings in pediatric populations: IGFBP1, IGFBP2 (markers of insulin sensitivity), and sex hormone binding globulin (SHBG), for which rapid early declines predict insulin resistance independently of adiposity, all aligned with our proteomic signals.^59^ ^60^ and downstream complications.^56^ In addition, proteins long implicated in adult CKMD—such as leptin (LEP), FABP4, and RARRES2—were recapitulated in children, suggesting shared molecular axes across lifespan.^61,62^ ^63^ In addition to well-established proteins, our analysis also revealed several proteins with plausible mechanistic roles but limited prior evidence in CKMD. These include BACE1, GPR158, FGFBP3, USP28, CRELD2, among others.^64,65^ ^66,67^ ^68^ ^69^ ^70^ Such proteins operate in signaling, cellular stress, or turnover pathways, and represent interesting candidates for future functional follow-up.

Pathway analysis across the CKMD proteome revealed associations with pathways of cholesterol metabolism, inflammation and hemostatic mechanisms, and metabolism (**Figure 2C-D**). Mapping to the Human Genetic Evidence (HuGE) framework demonstrated genetic support for many of the targets in the pediatric CKMD proteome with CKMD traits, including anthropometric, cardiovascular, hepatic, and renal phenotypes (**Figure 2E**).

**Figure 1:**
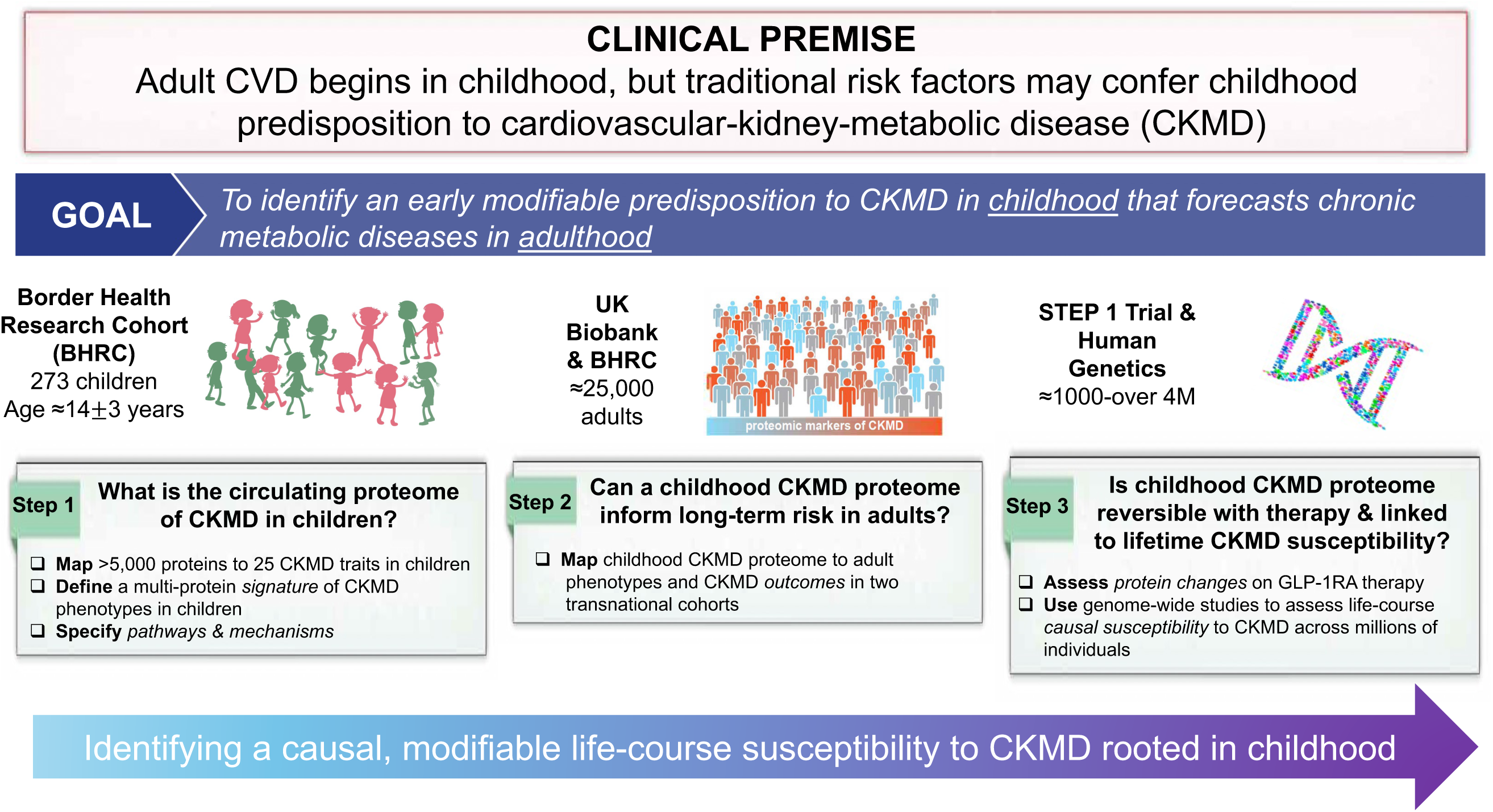
Study design.

**Figure 2:**
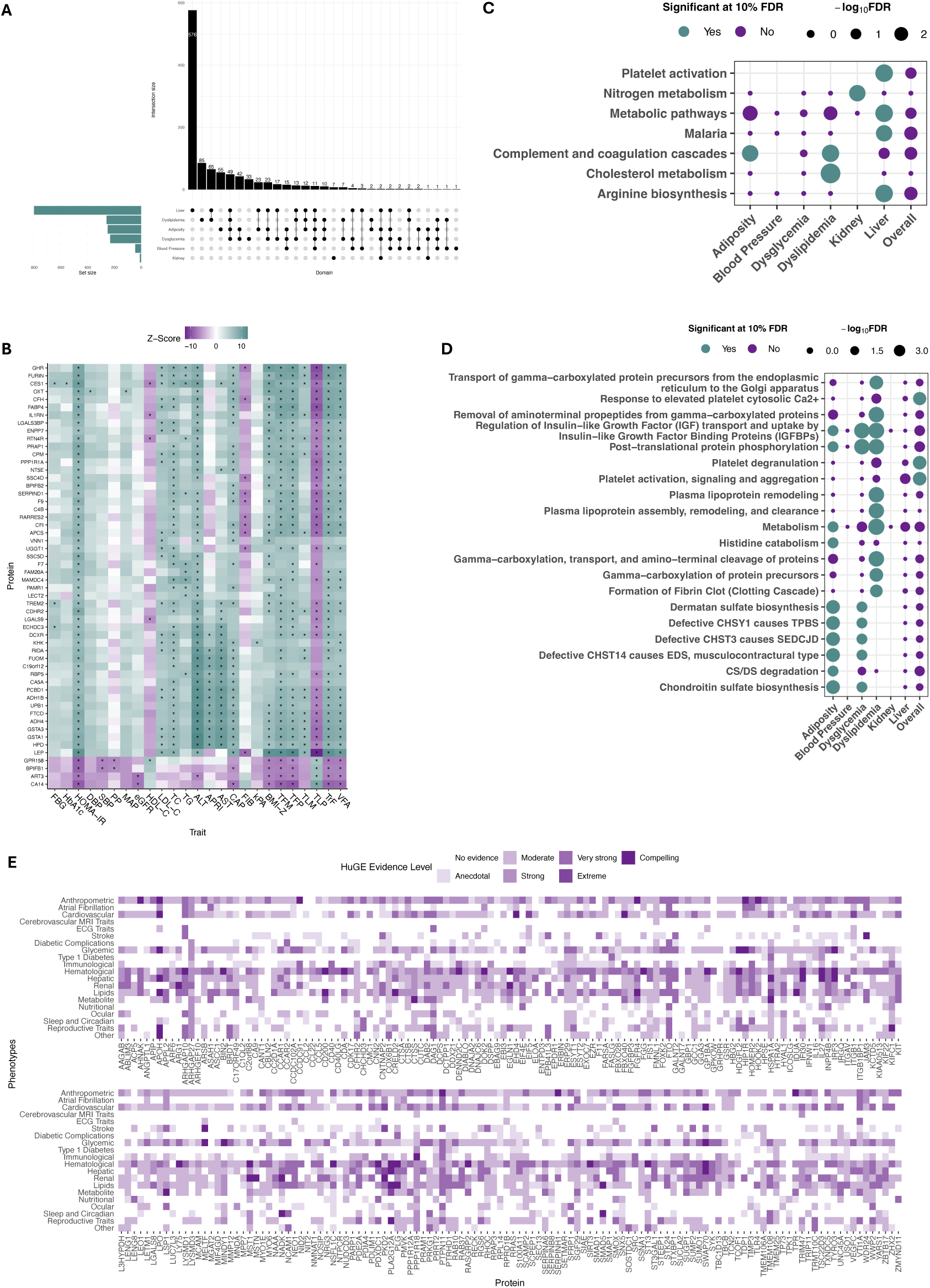
Proteomic architecture of CKMD in children. Panel (A) shows significant proteins across 6 domains (all at 5% FDR), demonstrating shared association. Panel (B) shows regression z-scores in linear mixed models (adjusted for age and sex) for proteins significant in at least four of the six phenotype domains. The * denotes significance at 5% FDR. Panels (C) and (D) show pathway analyses (KEGG and Reactome, respectively). Panel (E) denotes genetic evidence (HuGE) for select proteins from pediatric studies (as described in **Methods**).

### Characterizing the similarity between the pediatric and adult CKMD proteome

Understanding links between the serum proteome and CKMD phenotypes in children— specifically, whether these links recapitulate relationships classically observed in adults—is key to identifying nascent, modifiable features of CKMD. Accordingly, we next quantified the association between the circulating proteome and the 25 pediatric CKMD phenotypes across the six defined domains (**Supplemental Table 2**). We identified 2,916 protein-phenotype associations (at 5% FDR; the “CKMD proteome”; full results in **Supplemental Table 1A and 1B**). This represents 1,064 unique proteins, showing that a significant fraction of those proteins were significant across pediatric CKMD phenotypes (**Figure 2A**), and 65 were consistently associated with at least four CKMD domains (**Figure 2B**). While a complete mechanistic ontology lies beyond the scope of this work, the pediatric CKMD proteome (see curated entries in **Table 2**) reveals coherent signals that align with key metabolic processes in CKMD.

**Table 2.** Selected functional curation of proteins across multiple domains. This list includes proteins related to 5 domains (ACY1, ALDH1A1, CHGB, CKB, GPD1, GUSB, IGFBP1/2, IGSF9, C-peptide, SHBG) as well as additional selected proteins (as examples) with prior evidence in CKMD.

| Protein | Example domain | Direction (+ indicates higher protein ~ more adverse) | Function and prior studies in children (where reported) |
| --- | --- | --- | --- |
| **C-peptide** | Dysglycemia | + | Marker of beta-cell function and reserve, with higher levels suggesting hyperinsulinemia; increased in children with impaired glucose tolerance after glycemic challenge<sup>10</sup> |
| **Immunoglobulin Superfamily Member 9 (IGSF9)** | Liver | + | Immunoglobulin superfamily 9 involved in cell adhesion; related to hepatic steatosis and fibrosis in adults with HIV<sup>106</sup> |
| **Glycerol-3-phosphate dehydrogenase 1 (GPD1)** | Liver | + | Glycerol-3-phosphate dehydrogenase 1, involved in carbohydrate metabolism in the liver; mixed mechanistic findings, with congenital deficiency linked to hypertriglyceridemia<sup>107</sup> and liver disease,<sup>108</sup> while expression increased in adipose tissue in adults with obesity and associated with BMI;<sup>50,51</sup> expression responsive to weight loss intervention<sup>55</sup> |
| **Aminoacylase 1 (ACY1)** | Liver | + | Aminoacylase-1; high liver expression, levels in adults related to diabetes and hepatic steatosis<sup>49,53,54</sup> |
| **Chromogranin B (CHGB)** | Dysglycemia | - | Chromogranin B; involved in secretion of catecholamines (nervous system) and in insulin granule trafficking<sup>44</sup> (pancreatic beta-cells); genetic studies of *CHGB* variation and CHGB-null mice suggest lower levels related to higher blood pressure<sup>45</sup>; previous association with metabolic traits in adults<sup>46</sup> |
| **Aldehyde Dehydrogenase 1 Family Member A1 (ALDH1A1)** | Liver | + | Aldehyde dehydrogenase; important in metabolic processing of aldehydes in the liver; expression in pediatric livers lower than adults<sup>109</sup>; increased expression may protect against hepatic oxidative stress/inflammation during liver injury<sup>52</sup> |
| **Creatine Kinase B (CKB)** | Adiposity | - | Creatine kinase B; involved in high-energy phosphate transfer in highly metabolic tissues; increase in endoplasmic reticular stress during obesity epigenetically *reduces CKB* expression, increasing inflammatory chemokine expression in fat;<sup>57</sup> reduction in CKB in mice impairs thermogenesis and increases obesity<sup>58</sup> |
| **Beta-glucuronidase (GUSB)** | Dysglycemia | + | Beta glucuronidase; prior studies in adults link circulating levels to diabetes<sup>47</sup> |
| **Insulin-like growth factor-binding protein (IGFBP1/2)** | Dysglycemia, adiposity | - | Insulin-like growth-factor binding proteins; linked to lower risk of hepatic steatosis, obesity, atherosclerotic heart disease, and insulin resistance<sup>48,49</sup>; previous evidence in children suggests IGFBP1 is an early marker of insulin resistance<sup>59</sup> |
| **Sex hormone-binding globulin (SHBG)** | Dysglycemia, blood pressure | - | Sex hormone binding globulin; involved in sex hormone metabolism; early reduction in SHBG related to faster decline in insulin sensitivity independent of weight<sup>60</sup>; levels lower in obesity, reversible with weight loss, with higher levels linked to hepatic steatosis and widespread metabolic dysfunction<sup>56</sup> |
| **β-site amyloid precursor protein cleaving enzyme 1 (BACE-1)** | Dysglycemia | - | Overexpression can lead to increased hepatic insulin resistance in murine models, potentially via BACE1-mediated cleavage of the insulin receptor; pharmacologic targeting of BACE1 can abrogate these effects<sup>64,65</sup> |
| **G-protein coupled receptor 158 (GPR158)** | Dysglycemia | - | Involved in CNS-based regulation of energy metabolism and nutrient intake;<sup>66</sup> variants in GPR158 may impact liability to weight gain<sup>67</sup> |
| **Guanylate binding protein 1 (GBP1)** | Liver | + | Involved in pyroptosis in inflammatory liver injury<sup>110</sup> |
| **Fibroblast Growth Factor Binding Protein 3 (FGFBP3)** | Dyslipidemia | + | FGFBP3-null mice have lower lipid and triglyceride levels, and supplementation of FGFBP3 in obese mice can ameliorate metabolic dysfunction, weight gain, and liver steatosis; mechanism via interaction with FGFs<sup>68</sup> |
| **Ubiquitin specific peptidase 28 (USP28)** | Liver | + | High expression in human metabolic-associated steatotic liver disease and steatohepatitis; functions via stabilizing PPAR-gamma via blocking ubiquitination; lack of USP28 may lead to reduction in PPARγ signaling, and reducing hepatic inflammation<sup>69</sup> |
| **Cysteine-rich with EGF-like domains 2 (CRELD2)** | Liver | - | May limit ER stress and resilience; lack of CRELD2 may augment unfolded protein response and contribute to MASLD and changes in energy expenditure; may even have sex-specific effects<sup>70</sup> |
| **Fibroblast growth factor receptor 4 (FGFR4)** | Dysglycemia | - | Divergent results; FGFR4 modulates insulin sensitivity in murine obesity models<sup>111,112</sup> |
| **Cathepsin B (CTSB)** | Adiposity | - | Lysosomal protease; increased in white adipose tissue in mice; may regulate perilipin-1, which regulates lipolysis in fat<sup>113</sup> |
| **Transmembrane Protein 108 (TMEM108)** | Adiposity | - | Traditionally implicated in psychiatric disorders; TMEM108 mutant mice have poorer insulin sensitivity and shift metabolism toward glucose (away from lipid); metformin can rescue this effect<sup>114</sup> |

Across 685 adults and up to 25 phenotypes measured in both children and adults (**Table 1**), we observed a broad concordance in the magnitude and directionality of the phenotype-protein relationships (**Figure 3, Supplemental Table 3**). Of the 2,916 significant protein-phenotype associations, 1,883 (65%) were significant in adults with directionally consistent effects. Across domains, we observed the strongest pediatric-adult correlation in adiposity (r=0.61) and liver domains (r=0.69). While many proteins displayed expected associations with phenotypic data (e.g., ACY1, GOT1 with hepatic steatosis and fibrosis), we also observed “off-axis” proteins—those with divergent effect direction in children and adults—some of which have demonstrated pathophysiologic roles in CKMD (e.g., PON3,^71^ APOF^72^ for some dysglycemia phenotypes, including HbA1c) as well as those with no prior evidence in CKMD (e.g., MENT, NXPH1). Collectively, these results supported our hypothesis that a CKMD proteome— mechanistically relevant and classically described in adults—is identifiable in children years or decades before the onset of clinical consequences.

**Figure 3.**
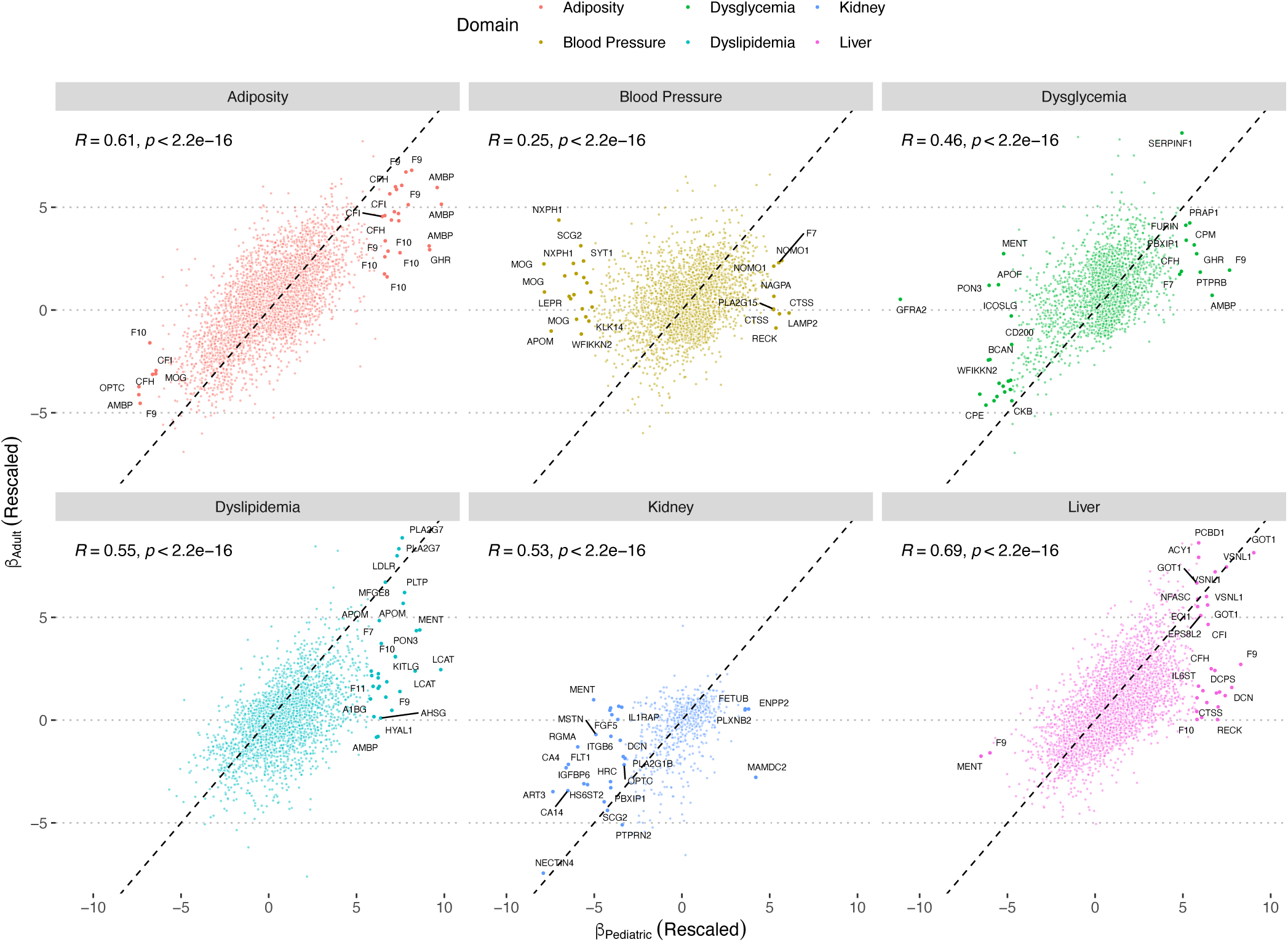
Concordance of CKMD phenome-proteome relationships between adults and youth. This figure depicts the correlation of pediatric and adult regression model effect sizes. The x-axis displays the rescaled beta coefficients for the pediatric results, and the y-axis shows the rescaled beta coefficients for the adult batch 2 results. Protein labeling is arbitrary.

### Generating a pediatric proteomic signature of CKMD

While the biological plausibility and concordance of proteome-CKMD phenome associations across adults and children suggest an early nascent phase in CKMD, identifying clear links to clinical risk factors or outcomes remains difficult, given the extended time horizon from childhood needed to capture clinical events. Therefore, we next constructed multi-protein-based signatures of CKMD phenotypes in children to test their relevance for <u>clinical</u> CKMD endpoints in adults. We used PCA to summarize 16 phenotypes measured in both children and adults, retaining 6 components (PCs) explaining ≈77% overall variation in the measured pediatric phenome (**Supplemental Figure 2**). For interpretability, we labeled PCs by clinical interpretation of features with high loadings, recognizing that each PC was loaded heterogeneously on CKMD phenotypes (**Figure 4**; loadings in **Supplemental Table 4**): (1) <u>pro-inflammatory</u> adiposity (PC1: primarily loaded on pro-atherogenic lipids, HOMA-IR, and BMI); (2) <u>liver fat/fibrosis</u> (PC2: loaded on transaminases and APRI); (3) <u>cholesterol</u> (PC3: loaded on LDL and total cholesterol); (4) <u>blood pressure</u> (PC4): loaded on SBP and DBP); (5) <u>kidney function</u> (PC5: loaded on eGFR and liver fibrosis); (6) <u>insulin resistance</u> (PC6, loaded on HbA1c, HOMA-IR, and fasting glucose). The clinical interpretation of “higher” PC values varied depending on loading directionality, with <u>higher</u> values for all PCs but PC3 representing a more <u>adverse</u> CKMD state. We generated proteomic signatures of each PC in LASSO regression to serve as transportable molecular “scores” of these summary traits by PC. The fit of our LASSO models (and number of proteins per model) is shown in **Figure 5** (**Supplemental Table 5** for model coefficients; mean cross-validated R^2^ 0.07-0.56 across PCs).

**Figure 4:**
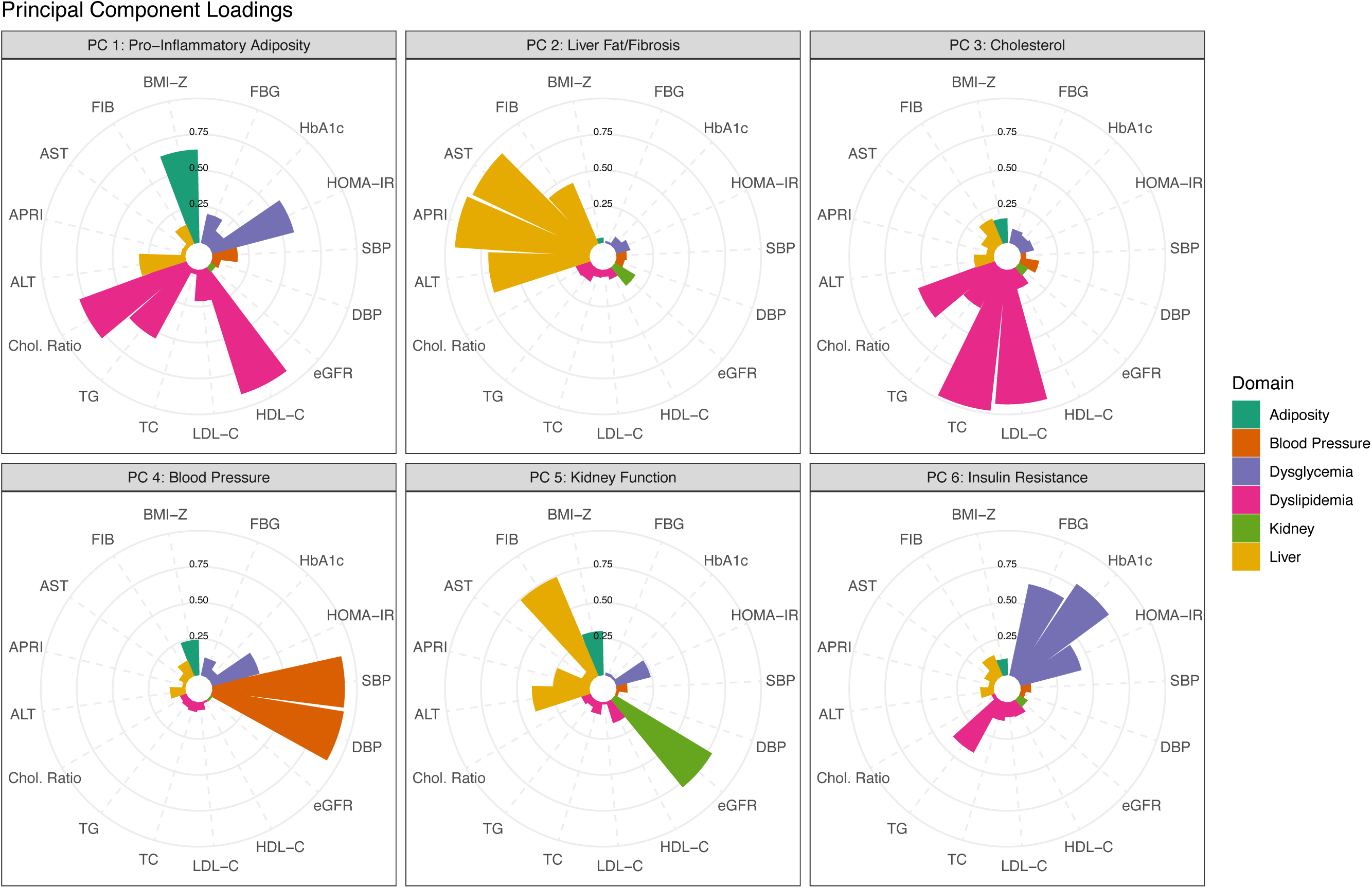
Principal components (PCs) of the pediatric CKMD phenome. These circular bar plots indicate the magnitude of the loadings per trait for each principal component of the pediatric phenotype matrix. The bars are colored by the domain of each trait. Abbreviations as in **Table 1**. “Chol.Ratio” is defined as the ratio of TC to HDL-C.

**Figure 5:**
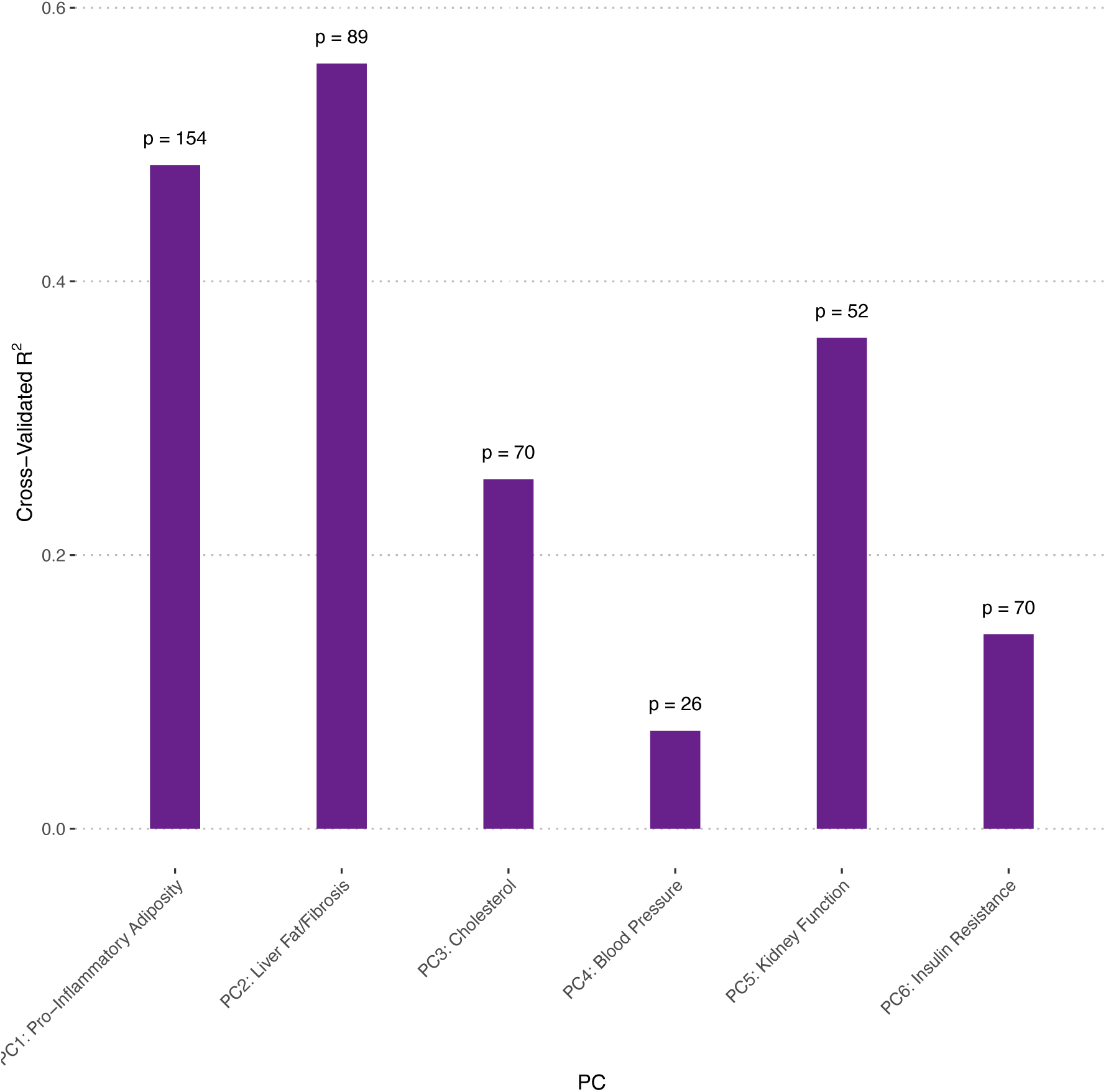
Goodness-of-fit for penalized regression models describing CKMD phenome in children. The R^2^ for each PC’s LASSO model is displayed alongside the number of proteins (p) that were used to fit those models.

### Applying pediatric CKMD proteomic signatures to adults identifies individuals with long-term susceptibility to cardiovascular and cardiometabolic disease in adulthood

We next examined whether proteomic signatures of pediatric CKMD would be associated with CKMD in adults. We first conducted a cross-sectional analysis in 494 adults from the same community, applying the pediatric proteomic PC scores to adult proteomic data and testing associations with CKMD-associated measures. This analysis demonstrated that adult associations of proteomic signatures with CKMD phenotypes were consistent with those observed in children (majority as expected based on PC loadings in children; **Figure 6**).

**Figure 6:**
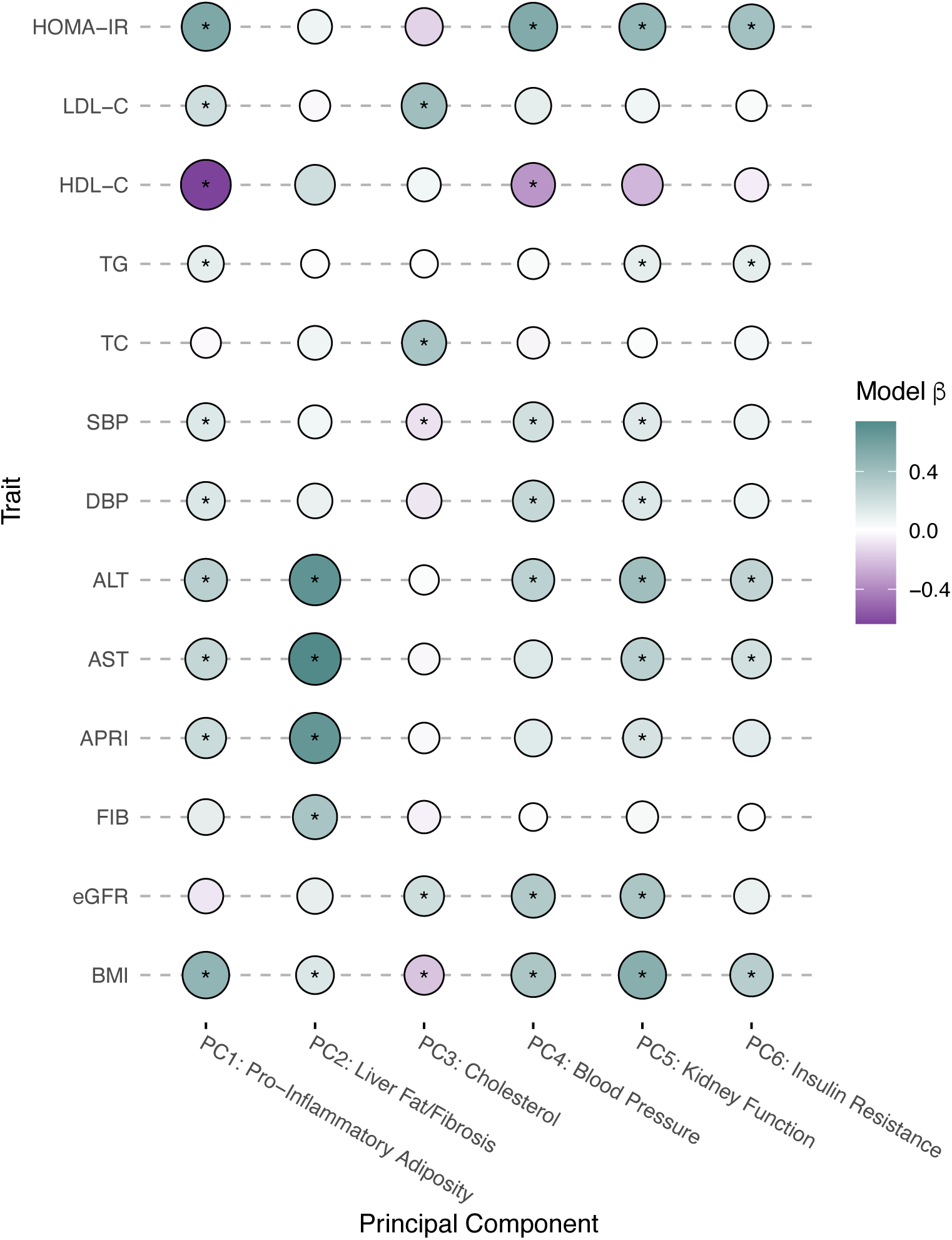
Mapping proteomic signatures of the pediatric CKMD proteome to adults. Age- and sex-adjusted association of each proteomic score (built on principal components of the <u>pediatric</u> CKMD phenome) in up to 521 Hispanic/Latino adults from the same population. Size and color intensity represent the magnitude of the effect size. The * indicates significance at P < 0.05. Correlations are Pearson.

In addition, we measured association between these pediatric CKMD signatures and incident CKMD-related outcomes in ≈25,000 individuals in the UK Biobank (characteristics in **Table 3**; median follow-up 13.7 years, 25^th^-75^th^ percentile 13.0-14.5 years for mortality). Pro-inflammatory adiposity (PC1), liver fat/fibrosis (PC2), and insulin resistance (PC6)—for which <u>higher</u> values in childhood signified <u>more advanced pediatric CKMD</u>—were associated with higher risk of all-cause mortality, diabetes, multiple cardiovascular diseases, sleep apnea, and fatty liver disease (**Figure 7**). Blood pressure (PC4) and kidney function (PC5)—also directly associated with more advanced pediatric CKMD—were linked to higher T2D risk, with heterogeneous associations with other conditions. In contrast, cholesterol (PC3), for which higher values in childhood were <u>less adverse</u>, had protective associations with cardiovascular and metabolic conditions in adults. Most of our observed associations were robust to comprehensive adjustment in adults (**Supplemental Table 6**). Collectively, these findings underscore the potential influence of early CKMD risk, as reflected in the childhood proteome, on the development of adult cardiovascular and metabolic disease.

**Figure 7:**
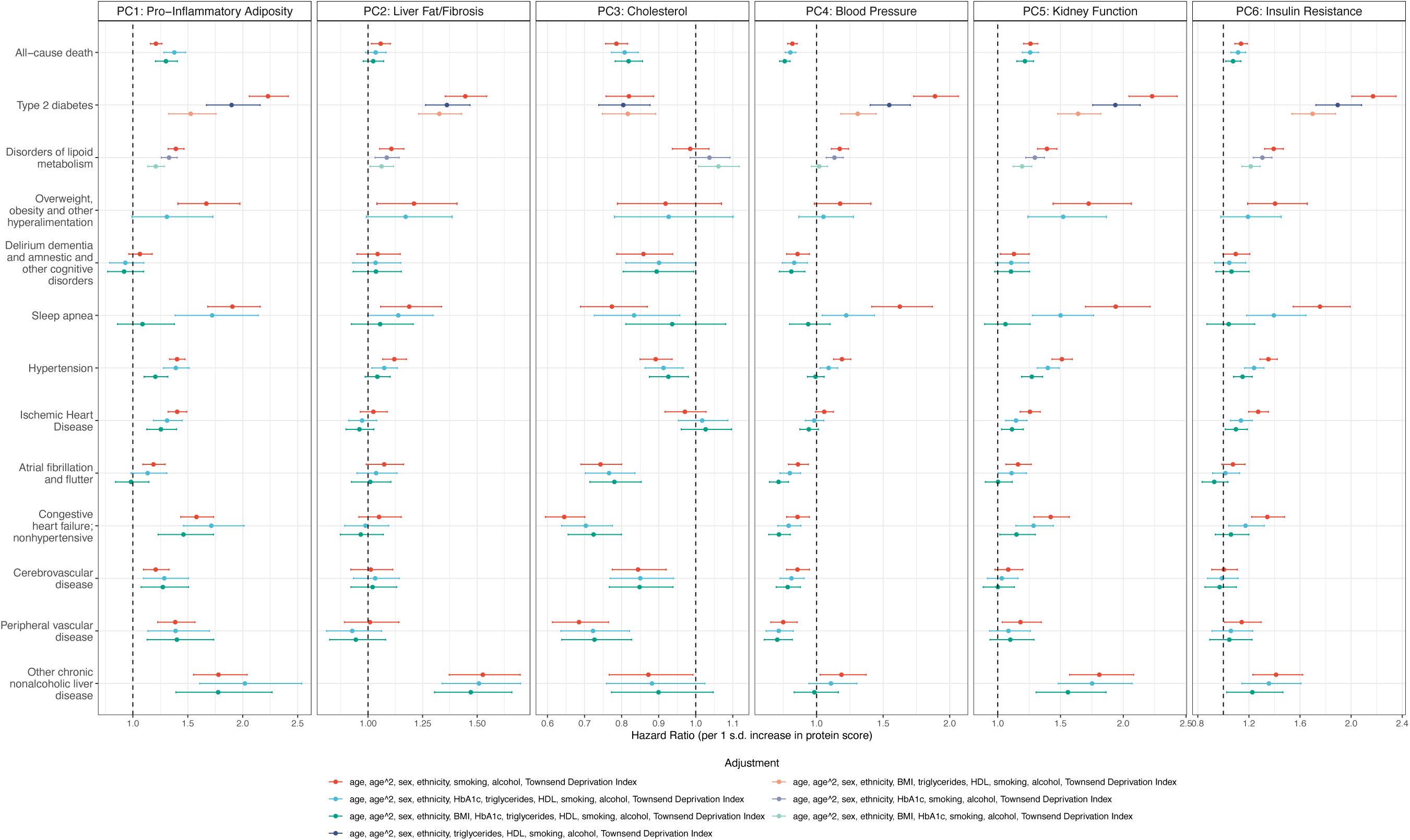
Long-term implication of the pediatric CKMD proteome on adult clinical risk. The forest plot depicts the hazard ratio (Cox models) of childhood-derived proteomic scores for CKMD-related endpoints in up to ≈25,000 individuals in the UK Biobank. The color of the marks in the plot indicates for which variables each model was adjusted. The error bars represent 95% confidence intervals of the hazard ratio.

**Table 3:** Demographic summary for adult participants analyzed from the UK Biobank. Models for each protein score included a subset of participants with complete data required to generate the respective protein score. The number of participants for each model is as follows: PC1 = 22,390; PC2 = 22,608; PC3 = 24,505; PC4 = 25,019; PC5 = 22,815; PC6 = 21,956. Fat mass and fat-free mass were measured by bioimpedance. Continuous variables are reported as mean (25^th^ percentile, 75^th^ percentile), and categorical variables are reported as N (%).

| Characteristic | N | Value |
| --- | --- | --- |
| Age (years) | 28,257 | 58 (50, 64) |
| Female | 28,257 | 15,273 (54%) |
| Ethnic Background | 28,257 |  |
| Asian or Asian British |  | 596 (2.1%) |
| Black or Black British |  | 690 (2.4%) |
| Mixed |  | 189 (0.7%) |
| Unknown \| Other |  | 495 (1.8%) |
| White |  | 26,287 (93%) |
| Body mass index (kg/m<sup>2</sup>) | 28,109 | 26.7 (24.1, 29.9) |
| Hemoglobin A1c (mmol/mol) | 26,770 | 35.3 (32.9, 38.1) |
| Triglycerides (mmol/L) | 26,840 | 1.47 (1.04, 2.14) |
| High-density lipoprotein (mmol/L) | 24,600 | 1.40 (1.16, 1.68) |
| Smoking status | 28,224 |  |
| Current |  | 3,067 (11%) |
| Never \| No answer |  | 15,389 (55%) |
| Previous |  | 9,768 (35%) |
| Alcohol use | 28,224 |  |
| Current |  | 25,754 (91%) |
| Never \| No answer |  | 1,371 (4.9%) |
| Previous |  | 1,099 (3.9%) |
| Townsend Deprivation Index | 28,219 | -2.0 (-3.6, 0.8) |
| Whole body fat mass (kg) | 27,602 | 23 (18, 30) |
| Whole body fat free mass (kg) | 27,652 | 51 (43, 62) |

### Mutability of the pediatric CKMD proteome during GLP-1RA therapy in adults

A key biological feature of a causal-functional biomarker of disease is its modifiability during therapies that address the underlying disease susceptibility. To explore the reversibility of proteins implicated by pediatric CKMD under metabolic therapy, we mapped 1,032 proteins associated with any CKMD outcome to data from the STEP 1 clinical trials of semaglutide (see **Methods**)^15^. We identified a striking relationship between pediatric-CKMD phenotypes and dynamic changes with GLP1-RA: proteins linked to <u>more adverse CKMD in children</u> were more likely to <u>decrease</u> following semaglutide therapy, and vice versa (Fisher’s exact test P < 2.2x10^-16^; **Figure 8A**). Notable examples included LEP, FABP4, CES1, and ACY1—all associated with greater pediatric insulin resistance, adiposity, and liver fat/fibrosis and with plausible CKMD-relevant mechanisms in adults (all reduced on GLP-1RA). Conversely, SHBG, WFIKKN2^53^, and IGFBP-2 represent examples of proteins with generally opposing effects relative to leptin in the pediatric sample and increased after GLP-1RA. In addition to putative mediators of CKMD risk, this approach also identified proteins with limited prior implication in diabetes or CVD, though with modifiability and relation to early CKMD (e.g., BCAN, ART3). A heatmap of the top 10 proteins (ranked by effect size) for each phenotype in the pediatric cohort is shown in **Figure 8B**. Proteins implicated by the CKMD phenotypes in children that were mutable with GLP-1RA therapy appeared distributed across key tissues and cell types, with strong representation of hepatocytes, pancreatic cells, adipose tissue, and brain tissue (**Supplemental Figure 3**).

**Figure 8:**
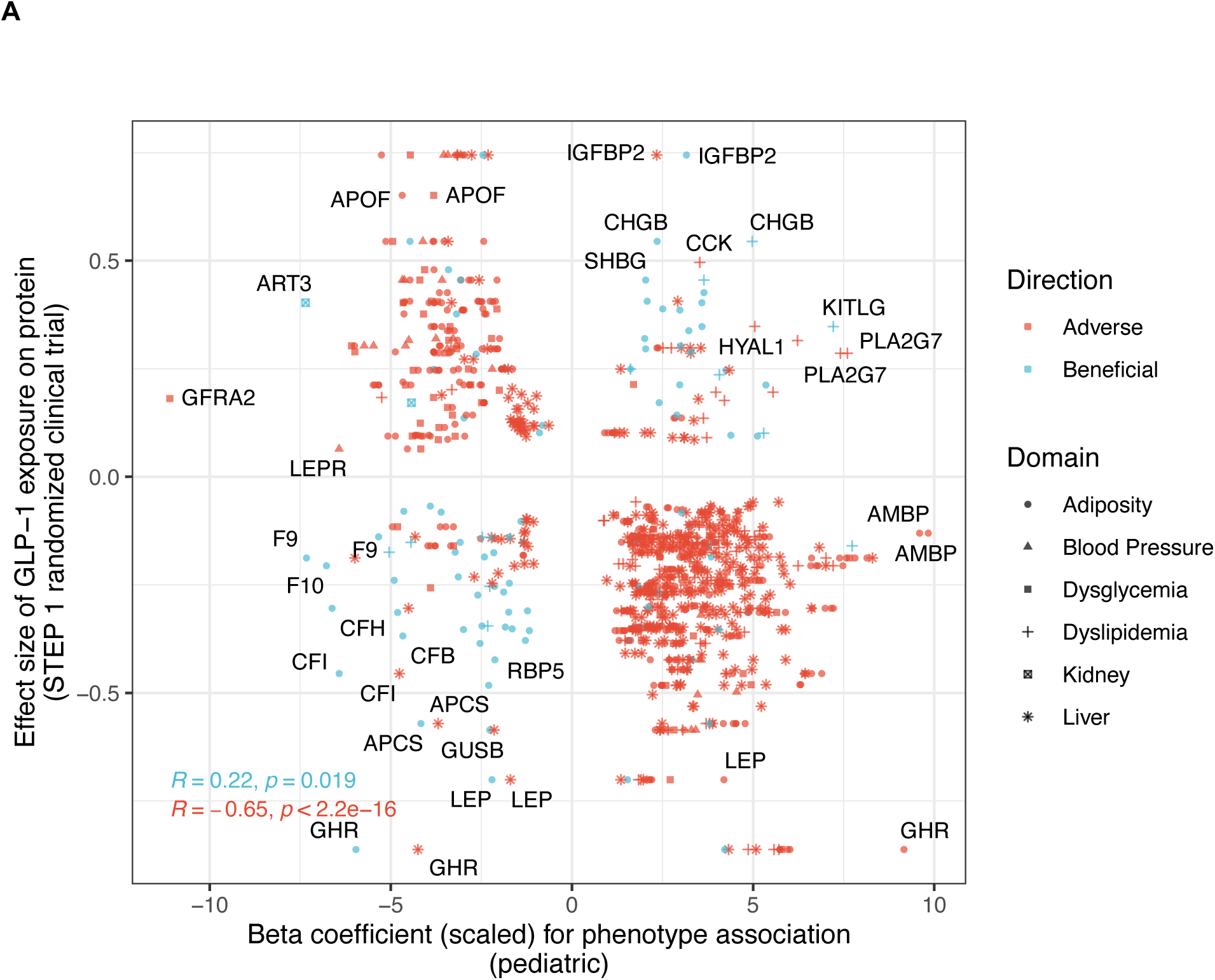
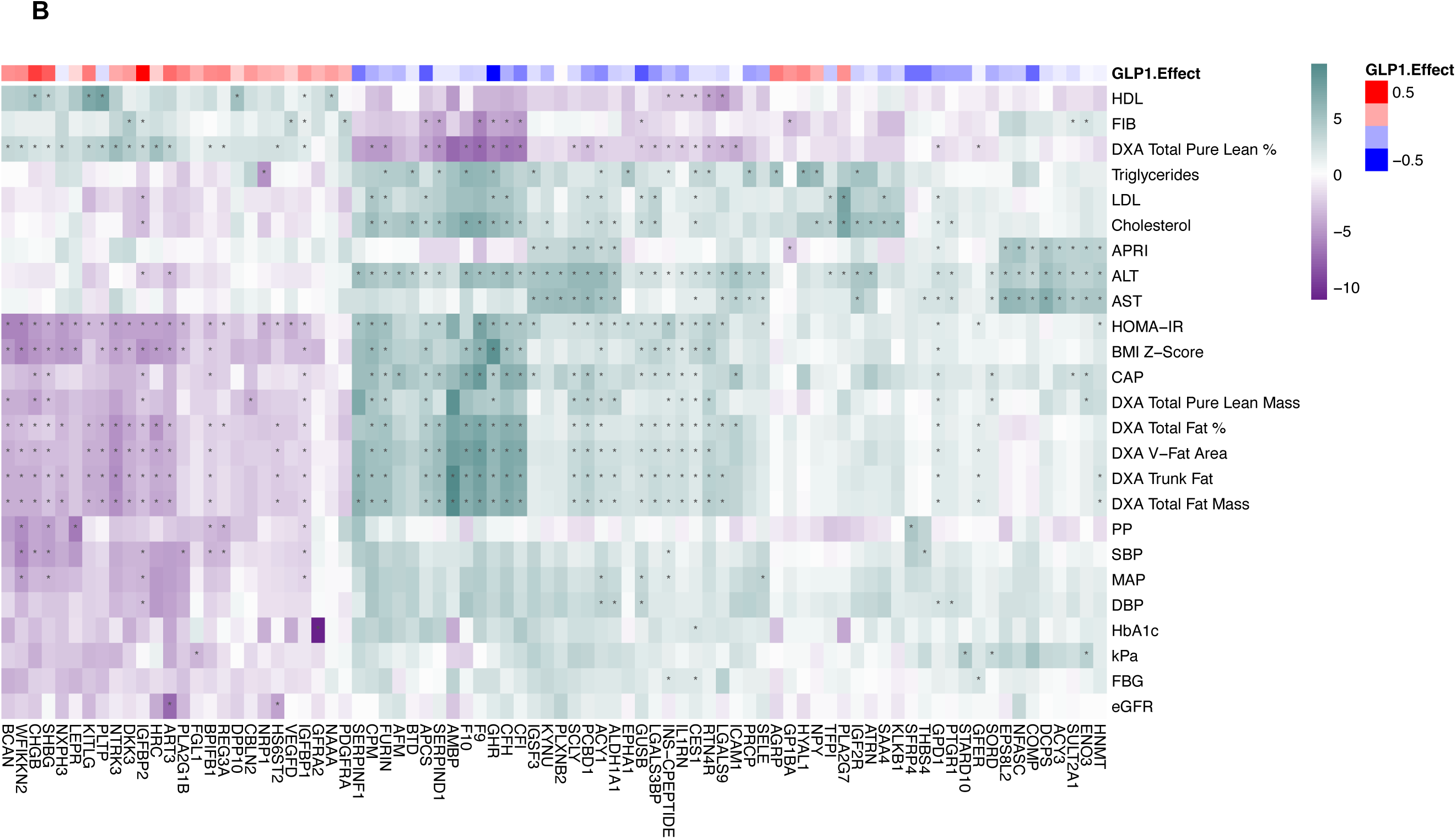
Dynamicity of the circulating pediatric CKMD proteome with GLP-1RA therapy. Panel (A) demonstrates the pediatric CKMD proteome across the 6 domains matched to proteomic data from the STEP1 clinical trial, showing the changes in circulating proteins after 68 weeks of therapy. Effect sizes were generated from linear regression models. Points are colored based on the directionality (adverse/beneficial) of the proteins’ relation with the corresponding CKMD phenotype (e.g., a protein with a positive coefficient for lean body mass is designated "beneficial," whereas a protein with a positive relation for BMI is designated "adverse"). Generally, we observed that proteins with "adverse" relations to CKMD phenotypes were decreased with GLP-1RA therapy and proteins with "beneficial" relations were increased. Panel (B) shows a visualization of the top 10 proteins (ranked by effect size) related to individual CKMD phenotypes in the pediatric cohort (with several proteins overlapping among phenotypes). Proteins that are positively associated with phenotypes (e.g., BMI, HOMA-IR) were generally reduced after GLP-1RA therapy.

### Genetic liability of the dynamic pediatric CKMD proteome is linked to lifetime CKMD

Genetic evidence linking the reversible pediatric CKMD proteome to adult CKMD clinical outcomes would provide support that lifetime exposure to these genetically regulated proteins influences CKMD trajectories. To that end, we performed a PWAS analysis using large GWAS (heart failure [N=1,946,349 individuals, 153,174 cases], BMI [N=419,163 in UK Biobank], diabetes [N=2,535,601 individuals, 428,452 cases], and renal function [creatinine; N=1,004,040]) to estimate the relationship between lifetime exposure to genetically regulated protein abundance and CKMD outcomes. We restricted analysis to 131 proteins that 1) were associated with pediatric CKMD phenotypes (at 5% FDR) and 2) had evidence of changing/improving after GLP-1RA (at 5% FDR). We observed strong associations with our reversible pediatric CKMD proteome across CKMD outcomes (**Figure 9**), including targets with cross-system, canonical physiologic roles with potential therapeutic relevance. Notably, we observed pediatric CKMD targets with potentially causal effects for BMI-specified mechanisms of energy homeostasis (e.g., SLIT2, via beige fat thermogenesis improvement of glucose sensitivity^73^), appetite (e.g., SCG3, via food intake regulation^74^), and adipose tissue inflammation and adipogenesis (SERPINE1/PAI-1^75^, COMP^76^). Importantly, pediatric CKMD targets also demonstrated potential causal effects on obesity-related downstream sequalae across biological pathways, including activin and TGF-beta signaling (INHBC^77,78^), matrix remodeling (TIMP4), oxidative homeostasis (GSTA1), autophagy (SPON2^79^), and lipoprotein metabolism (APOH, implicated in lipoprotein(a) metabolism^80^). In addition, we identified new targets not widely recognized in obesity, including HYAL1—significant across several outcomes in our PWAS analyses—involved in hyaluronan catabolism with potential roles in T2D and CVD^81^. Collectively, these results link lifelong genetic features to a proteome identified in children with downstream CKMD complications in adulthood.

**Figure 9:**
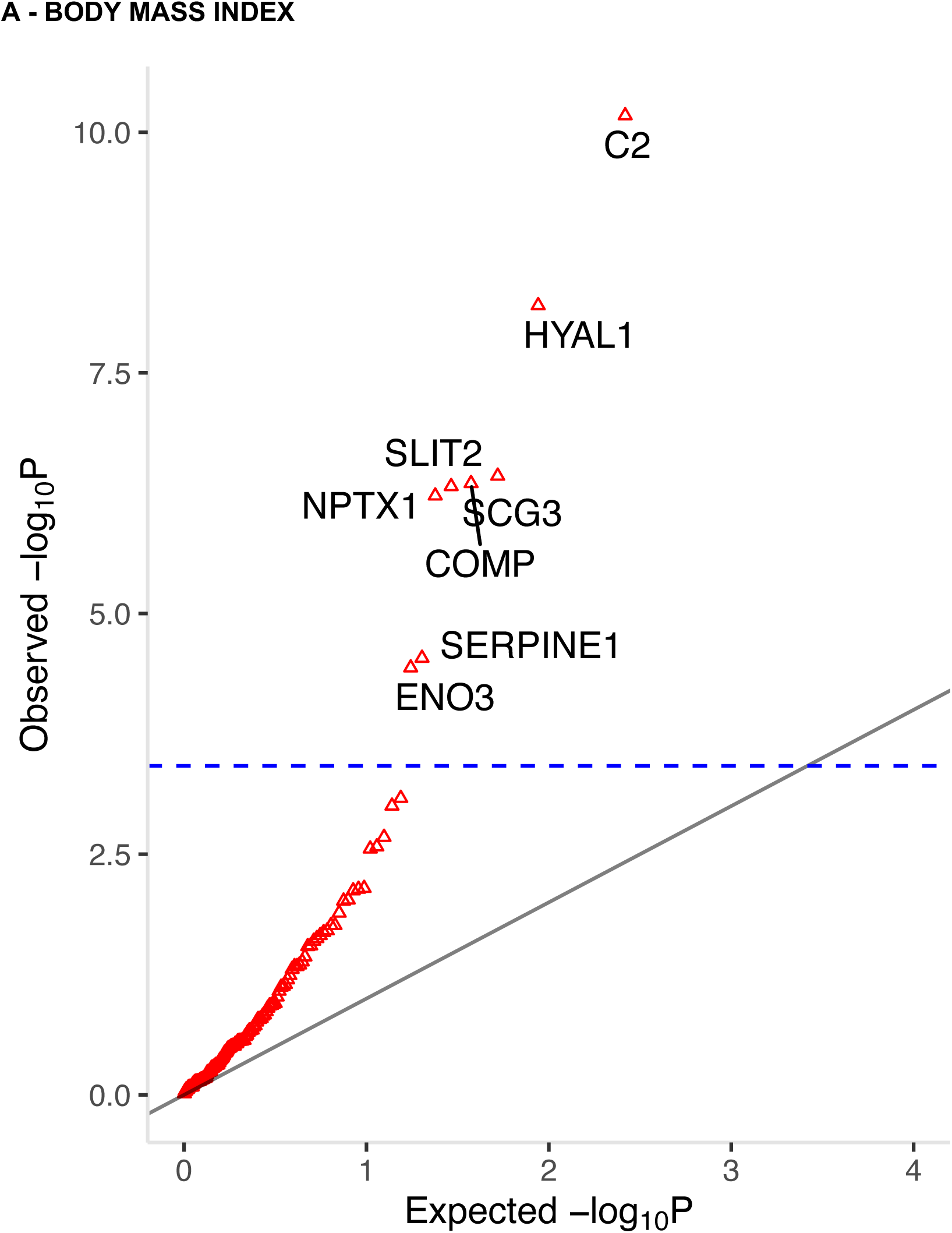
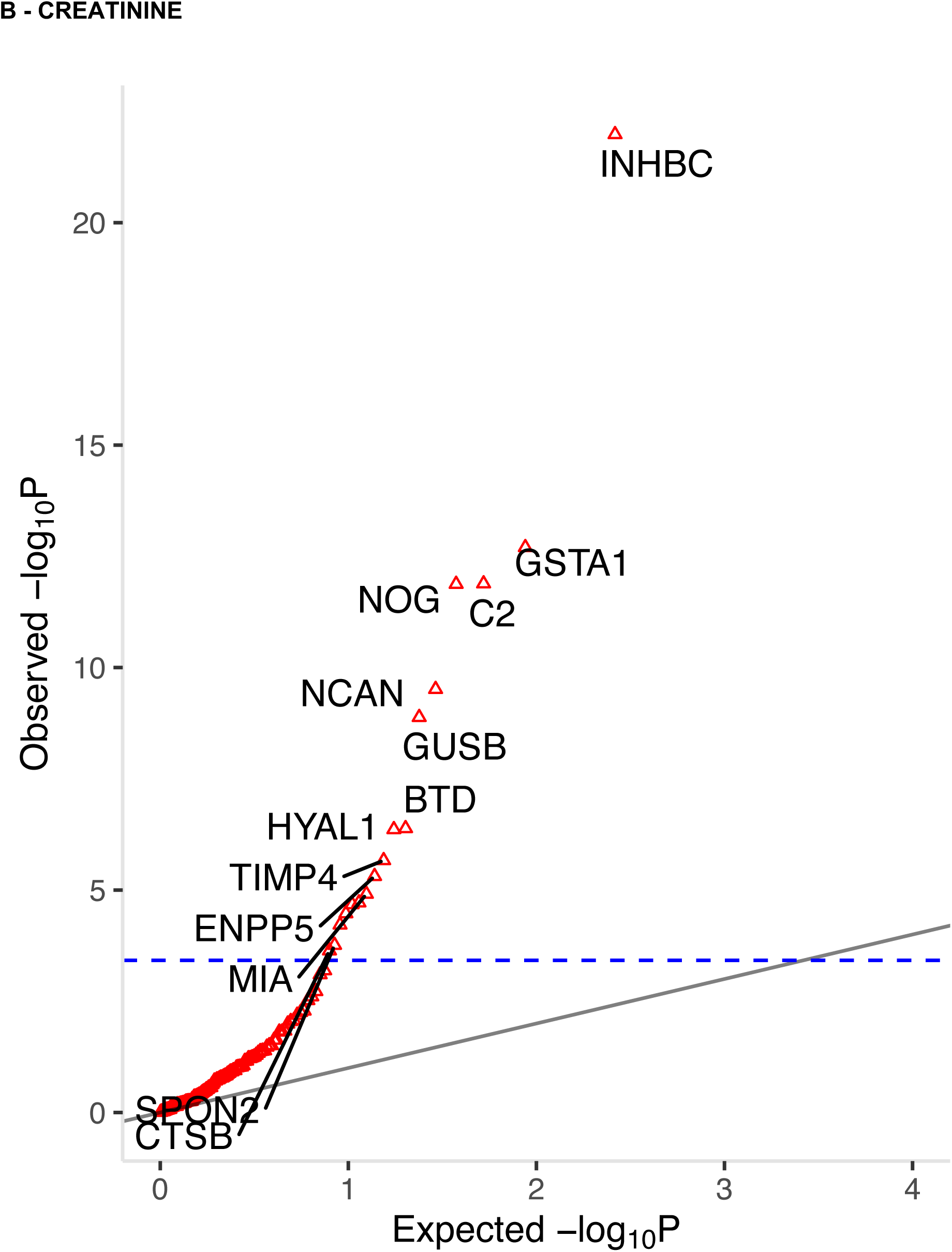
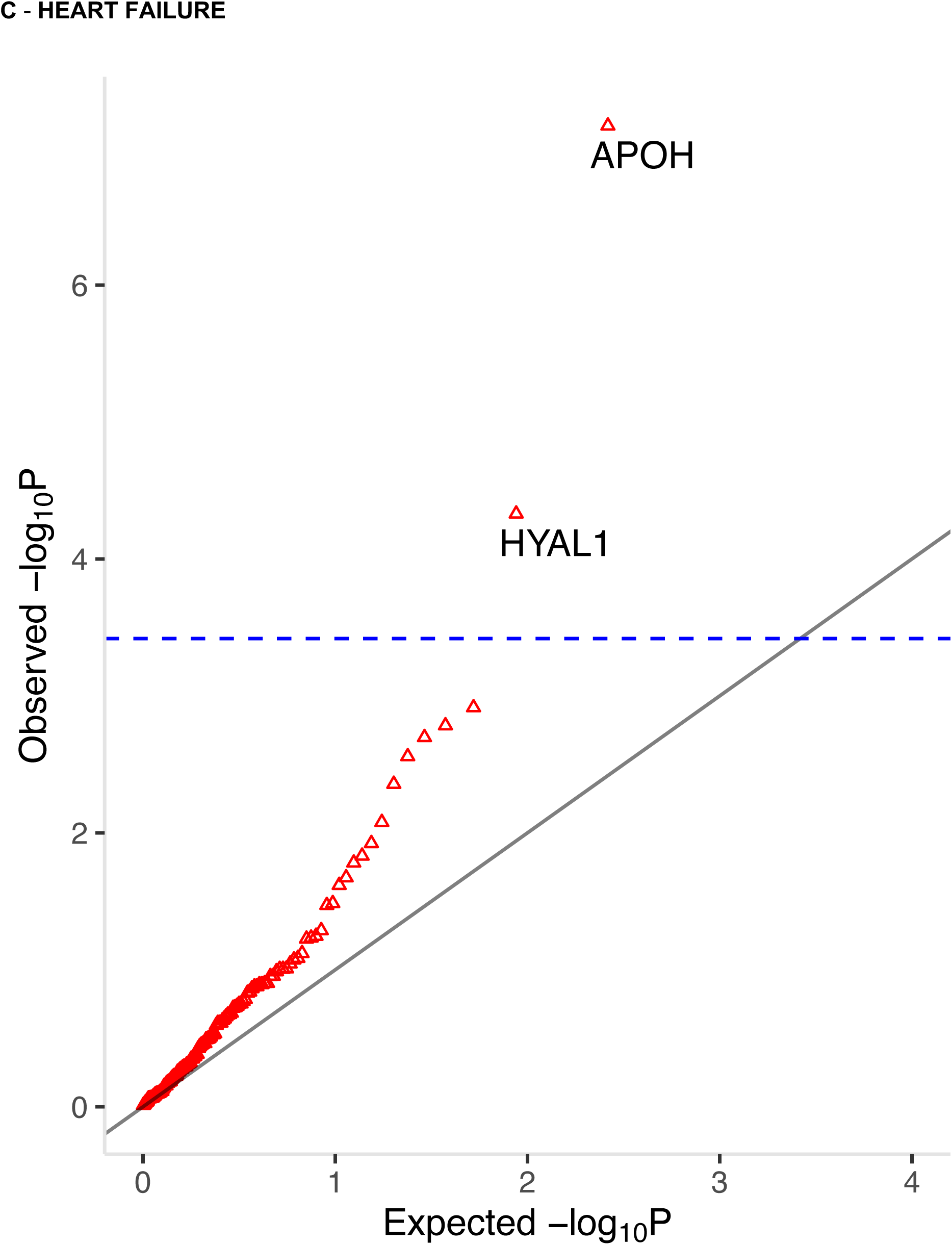
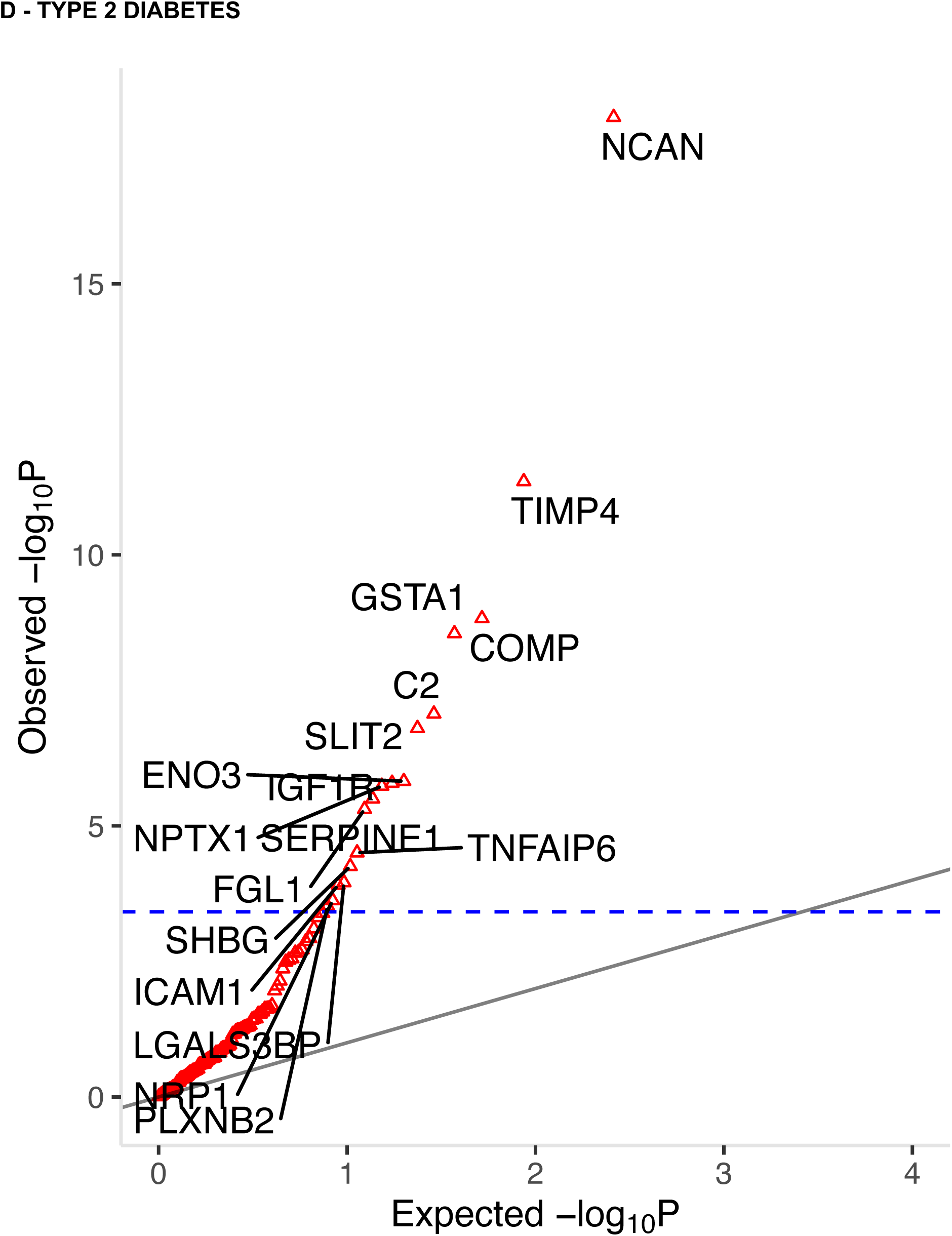
Genetic liability to a dynamic pediatric CKMD proteome is linked to long-term CKMD clinical outcomes. Each panel represents the results of proteome-wide genetic association studies (PWAS) for proteins associated with CKMD phenotypes in BHRC children and that were modifiable with GLP-1RA therapy in the STEP 1 trial adults. The blue dotted line represents a Bonferroni p-value threshold of 0.05 across the proteins tested. Proteins above this line (labeled) exhibit genetic evidence for association with the target outcome.

### Early developmental variation in a shared pediatric-adult CKMD proteome

How variable proteins linked to CKMD are during the adolescent-adulthood transition— and whether they reflect developmental age- or sex-related changes—is critical to positioning their use as potential reversible biomarkers of early CKMD. We leveraged a recently reported study of 100 individuals studied longitudinally from age 4 to 24 years in the Swedish BAMSE cohort^17^ to examine sources of variability in protein abundance. Focusing on proteins with evidence of significance (at 5% FDR) in children that replicated in BHRC adults, we observed broad variability in the CKMD-associated proteome (**Figure 10** for four select domains for visualization purposes; full abstracted results in **Supplemental Table 7**^17^). Overall, we observed a high degree of variation unexplained by age or sex for a large fraction of proteins associated with CKMD in children and adults (example trajectories for key metabolic proteins LEP, IGFBP2, ACY1, and CES1 in **Figure 10**). Age/sex explained a relatively low proportion of variability for our traits with the notable exception of BMI-Z score, which reflects growth and development. Of note, many proteins that displayed a substantial variability not explained by age or sex were modifiable through GLP1-RA therapy in adults (e.g., IGFBP1, IL1RN, TNFSF14, etc.). This substantial, modifiable residual variability in the CKMD-relevant proteome through early development suggests the potential for intervention to improve metabolic risk in children with implications for adult CKMD.

**Figure 10:**
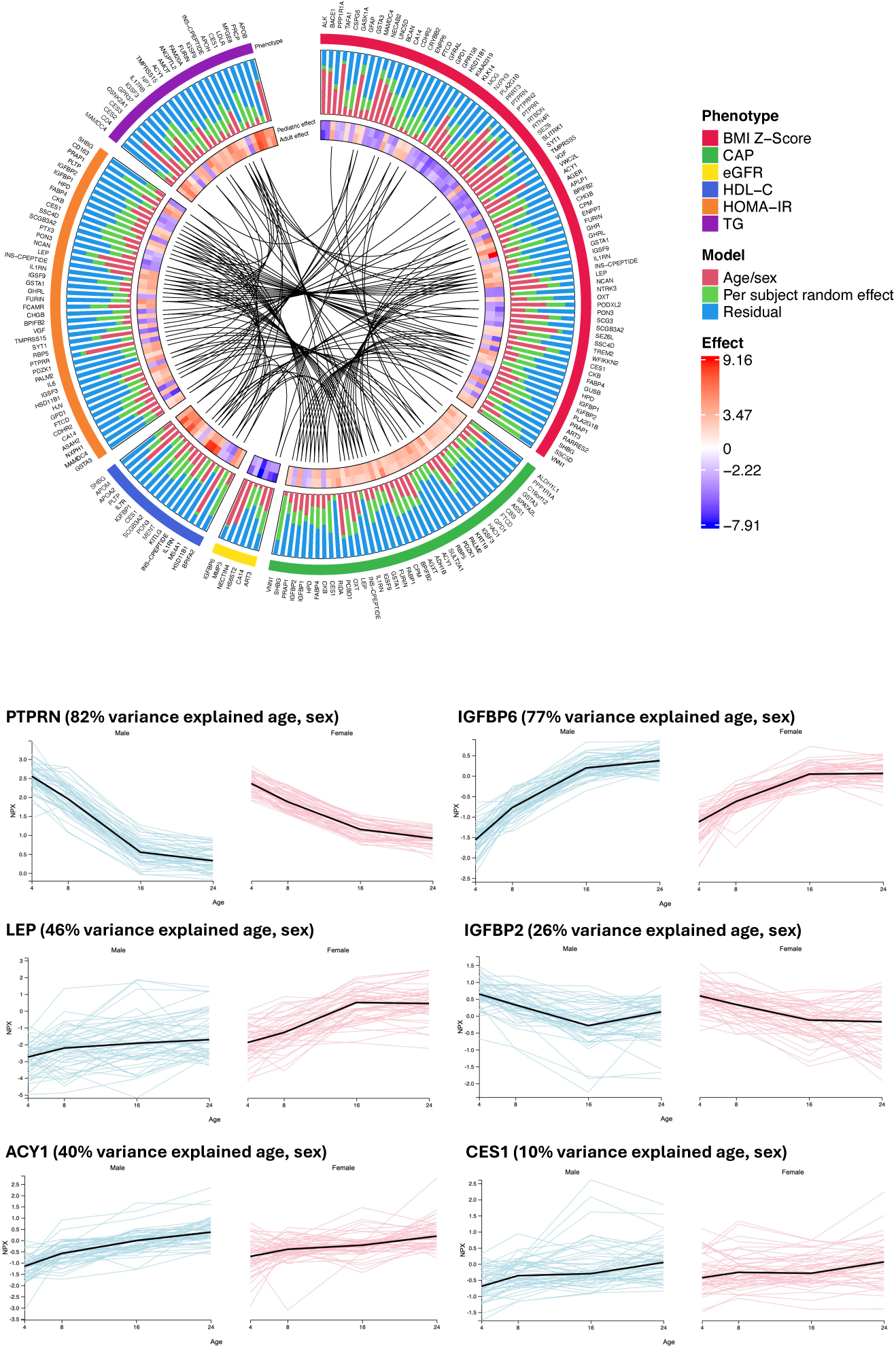
**Variation of the CKMD proteome during early development**. Top panel visualizes variance components from recent Bueno Alvez et al. (reference in text) for 6 phenotypes (BMI Z score, liver CAP, eGFR, HOMA-IR, triglyceride, and HDL). The outer bar plot shows the variance components (age/sex, per subject random effect, and residual. The inner layer shows the association effect size (beta coefficient) in BHRC children and adults. Inner lines link the same gene identified in different phenotypes. Regression coefficients for BHRC children and adults are scaled. The protein plots under the circus visualization depict protein levels (NPX) across early development to young adulthood from the Human Protein Atlas (see Methods). The top two proteins (PTPRN and IGFBP6) are included as examples of high variation explained by age and sex. Remaining proteins are examples across the spectrum of variance explained associated with CKMD.

## DISCUSSION

Increasing CKMD has stoked an alarming rise in early cardiovascular morbidity, prompting earlier, premature, and even “primordial” prevention to address life-course CVD risk.

Over the last four decades, evidence from early coronary artery pathology reports,^1^ studies in Bogalusa,^82^ and recent data in ≈36,000 children from the i3C Consortium,^5^ have implicated obesity and related CKMD phenotypes (hypertension, pro-atherogenic dyslipidemia, insulin resistance) as drivers of premature CVD. In response, pharmacologic approaches (e.g., pediatric GLP-1 receptor agonist therapy^35^) and lifestyle interventions (e.g., nutrition, physical activity) are increasingly emphasized to attenuate CKMD in childhood.

A critical gap in this effort is the ability to identify which children carry high future risk and to determine whether pediatric CKMD shares pathophysiologic continuity with adult CVD. CKMD screening in children is triggered by obesity prevalence, which often misses at-risk children^83^. Furthermore, clinical intervention thresholds for blood pressure and lipids in children are less stringent than those employed in adult care,^3,4^ and standard screening tests (e.g., fasting glucose) often miss a substantial amount of disease-defining risk in children.^9,10^ Although most at-risk children do not suffer from clinical cardiovascular events, CKMD imposes cumulative damage, with early exposure triggering potentially irreversible cascades toward CVD. While current pediatric therapies to prevent CVD have shown success,^35^ it is unclear whether pediatric CKMD reflects distinct biology or is simply an earlier manifestation of adult CVD.

Here, using integrative analysis across the pediatric CKMD phenome, circulating proteome, and human genome, we derived a pediatric CKMD proteome signature and found notable concordance with adult CKMD architecture. We also curated mechanistic evidence linking these proteins to adult disease (including CVD), suggesting that CKMD states in children mirror adult pathophysiology. When applied in adults, our multi-protein signature of composite CKMD phenotypes in children, reflecting potential shared underlying biological pathways across different phenotypes, was strongly associated with adult CKMD cross-sectionally and predicted incident clinical conditions over a decade. Importantly, many proteins linked to pediatric CKMD were modifiable with GLP-1RA therapy, with directionally consistent changes, and proteins linked to pediatric CKMD and responsive to GLP-1RA therapy exerted a potentially causal effect on adult CKMD outcomes revealed through genetics, implicating known and novel biological mechanisms with origins in childhood. Finally, using longitudinal data from children across ages 4-24 years, proteins linked to CKMD in adults and children generally exhibited substantial variation not explained by age/sex, suggesting an opportunity for intervention to improve metabolic risk at this early stage. Collectively, these results indicate that pediatric CKMD largely represents an early, reversible form of adult disease, highlighting opportunities for enhanced precision risk assessment and therapy starting in childhood, a critical point in CKMD development.

Although overt clinical manifestations of disease in childhood are still rare, early metabolic perturbations increase the life-course damage to the cardiovascular system that is likely important for long-term morbidity and mortality. For example, reduced physical activity and low physical fitness levels promote insulin resistance^84^ and subclinical atherosclerosis^85^ in children. Furthermore, pediatric CKMD is related to early myocardial tissue changes that presage heart failure^86^ as well as pro-atherogenic dyslipidemia and inflammatory biochemical states^82,87^ ultimately linked to adverse outcomes in adulthood.^5^ Given the limitations of standard clinical tests to capture this metabolic risk early (e.g., fasting labs), there is growing recognition that molecular biomarkers—“omics”—can detect risk decades before its onset and often independent of clinical risk factors^14,88–92^. However, despite great interest in the pediatrics field^93,94^, multi-omics analyses of CKMD in children remain sparse^95–98^. As an exception, Stratakis and colleagues used targeted proteomics (Luminex) in 471 children to identify inflammatory proteins, including IL-6, associated with advanced CKMD.^95^ Two broader proteomic studies have similarly linked proteins such as SHBG, LEP, ADH4, and ADH1A to central obesity and hepatic steatosis.^97,99^

Our results fundamentally extend these previous reports via several key innovations: (1) we surveyed a far broader proteomic space than previously quantified; (2) we focused on a high-risk pediatric population with a high prevalence of obesity; (3) we included direct, imaging-based measures of metabolic-associated steatotic liver disease (MASLD) , an early mediator of CKMD risk;^49^ and (4) we benchmarked pediatric proteomic signatures against adult outcomes and phenotypes, including adults both from the same community and from a geographically distinct population. Of note, the multi-system CKMD outcomes were most strongly associated with proteomic signatures of hepatic disease, a condition that affects nearly 10% of children^100^ and may serve as a harbinger of adult CVD.^101^ Moreover, beyond ascribing risk in children to long-term outcomes in adults, we observed strong consistency in phenome-proteome relationships across the life-course, consistent with observations in accelerated aging. Several proteins linked to pediatric CKMD mapped to known adult CKMD-relevant pathways likely operant across age, including beta-cell function and inflammation, as well as new molecular mechanisms of early CKMD biology. Finally, dynamic modulation under GLP-1RA therapy, genetic evidence supporting causal roles across the life-course, and early unexplained variation at the adolescent-adulthood transition provide strong support that these proteins mark phenotypic states in children that are biologically and therapeutically relevant.

This transferability of phenotype and outcome from childhood to adulthood has important clinical implications. Universal promotion of cardiovascular health in children and adolescents remains critical but has yielded limited success,^102,103^ yet our data suggest that we can and must redouble these efforts. Pediatric obesity is increasingly being managed pharmacologically and surgically (with a nearly 600% increase in pediatric GLP-1RA prescriptions between 2020-2023) which should be further evaluated.^104,105^ In a precision-based framework to prioritize youth at significant CKMD risk to receive therapies is important to balance benefits with therapeutic burden and risk. The observed reversibility and strong predictive capacity of the composite signatures in childhood to CKMD strongly support its potential for precision risk assessments in children. Nevertheless, our study has several limitations. The studied population was derived from single county in Texas and is of Hispanic ethnicity with admixed European and Native American ancestry, which may limit generalization to other populations. However, we observed consistent findings across populations (UK Biobank) minimizing this concern. In addition, while GLP-1RA did appear to impact proteins we identified here, we did not specifically examine the dynamicity of the proteome over time in the pediatric sample; future studies with adequate serial sampling are needed. The proteomic panels used were relative quantification (Olink Explore), and more parsimonious studies with absolute quantification are required for clinical translation. Importantly, unlike genomic studies requiring 10^3^-10^5^ participants, our proteomic approach achieved larger clinically relevant effect sizes with substantial power at a significantly sample size (albeit still large for deeply phenotyped pediatric populations), likely due to greater biologic proximity of the proteome to the measured phenotypes and outcomes.

In summary, we delineated the proteomic architecture of CKMD in 273 children across 25 phenotypes, demonstrating similarities between the CKMD phenome-proteome relation across age, its biological plausibility, relevance to long-term clinical CKMD risk in adults through large cohort observational studies and human genetics, and mutability with GLP-1RA therapy. Our results support the hypothesis that CKMD exists on an age continuum with proteomic signals relevant to long-term irreversible outcomes appearing early in life at a reversible stage. Given the substantial effect size of the pediatric CKMD proteome on adult clinical CKMD, these observations underscore the importance of precision assessment in children at this reversible stage, potentially via the circulating proteome, to improve CVD-free longevity.

## Data Availability

All deidentified data produced in the present study are available upon reasonable request to the authors.

## AUTHOR CONTRIBUTIONS

JML, HMH, ASP, QS, AL, ABP, SZ, WZ, XZ, VLB, EGF, RR, AS, EHF, MYA, AB, CAPB, ERG, PGL, JEB, KEN, and RVS contributed to data analysis. JML, HMH, JBM, SPFH, PGL, JEB, KEN, and RVS contributed to study conception and design. JML, HMH, ASP, QS, ERG, PG, JEB, KEN, and RVS contributed to writing the manuscript. JML, HMH, ASP, AGH, AL, ABP, SZ, WZ, XZ, VLB, EGF, RR, AS, JKS, TW, JT, AG, LEP, LF, HC, MK, MG, KAM, ML, KLY, QW, ERG, JBM, SPFH, PGL, JEB, KEN, and RVS edited the manuscript. All authors contributed to critical review of the manuscript. EHF, MN, ASP, BL, JEF, ERG, JBM, SPF, PG, JEB, KEN, and RVS contributed to data acquisition.

## ACKNOWLEGEMENTS, FUNDING, AND DISCLOSURES

The authors would like to thank the BHRC cohort team, particularly Rocío Uribe who recruited and interviewed the participants; Marcela Morris, BS, and her teams for laboratory support; Valley Baptist Medical Center in Brownsville, Texas, for providing space where our Center for Clinical and Translational Science Clinical Research Unit is located; and the community of Brownsville and the participants who so willingly contributed to this study in their city. This study was funded in part by Center for Clinical and Translational Sciences and a National Institutes of Health (NIH) Clinical and Translational Award (grant UL1TR000371) from the National Center for Advancing Translational Sciences. Funding for the BHRC is supported by the following NIH grants: U54AG089326, R01DK139598, R01HL142302, R01HL163262, R01DK127084, and U01CA288325. JML was supported by NIH grant T32HG008341. JML, HMH, JBM, SPFH, JEB, and KEN were supported by NIH grant U01CA288325. JML and ML were supported by R01AG078452. ASP was supported by NIH grant K23HL179316 and is a co-inventor on patents of molecular signatures of fitness, liver and lung disease. HMH, ABP, KAM, JBM, SPFH, PGL, JEB, and KEN were supported by NIH grant R01DK139598. ABP, JBM, SPFB, JEB, and KEN were supported by NIH grants R01HL142302 and R01DK122503. AB was supported by NIH grant T32GM145734. AG, JBM, SPFH, JEB, and KEN were supported by NIH grant R01DK127084. MN is supported by National Institutes of Health (R01HL156975, R01HL131029, and R21AG086679). MK was supported by NIH grant U24DK132715. JBM, SPFH, JEB, and KEN were supported by NIH grant U54AG089326. ERG is supported by National Institutes of Health (grants U01DK140952 and R01HG011138, P60DK020593) and a grant from the Vanderbilt Diabetes Center (Functional Multi-Omics of Diabetes and Metabolism Core). ERG has performed consulting for Thryv Therapeutics and has pending patents or disclosures on molecular biomarkers of fitness, cardiovascular diseases and phenotypes, and metabolic health, use of RNAs as therapeutics and diagnostic biomarkers in disease, and methods in metabolomics. RVS is supported by grants from the National Institutes of Health and institutional endowment funds. RVS has equity ownership in and is a consultant for Thryv Therapeutics and is a co-inventor on pending patents or disclosures on molecular biomarkers of fitness, lung disease, cardiovascular diseases and phenotypes, and metabolic health, use of RNAs (including spatial) as therapeutics and diagnostic biomarkers in disease, and methods in metabolomics.

---

**Supplemental Figure 1.**
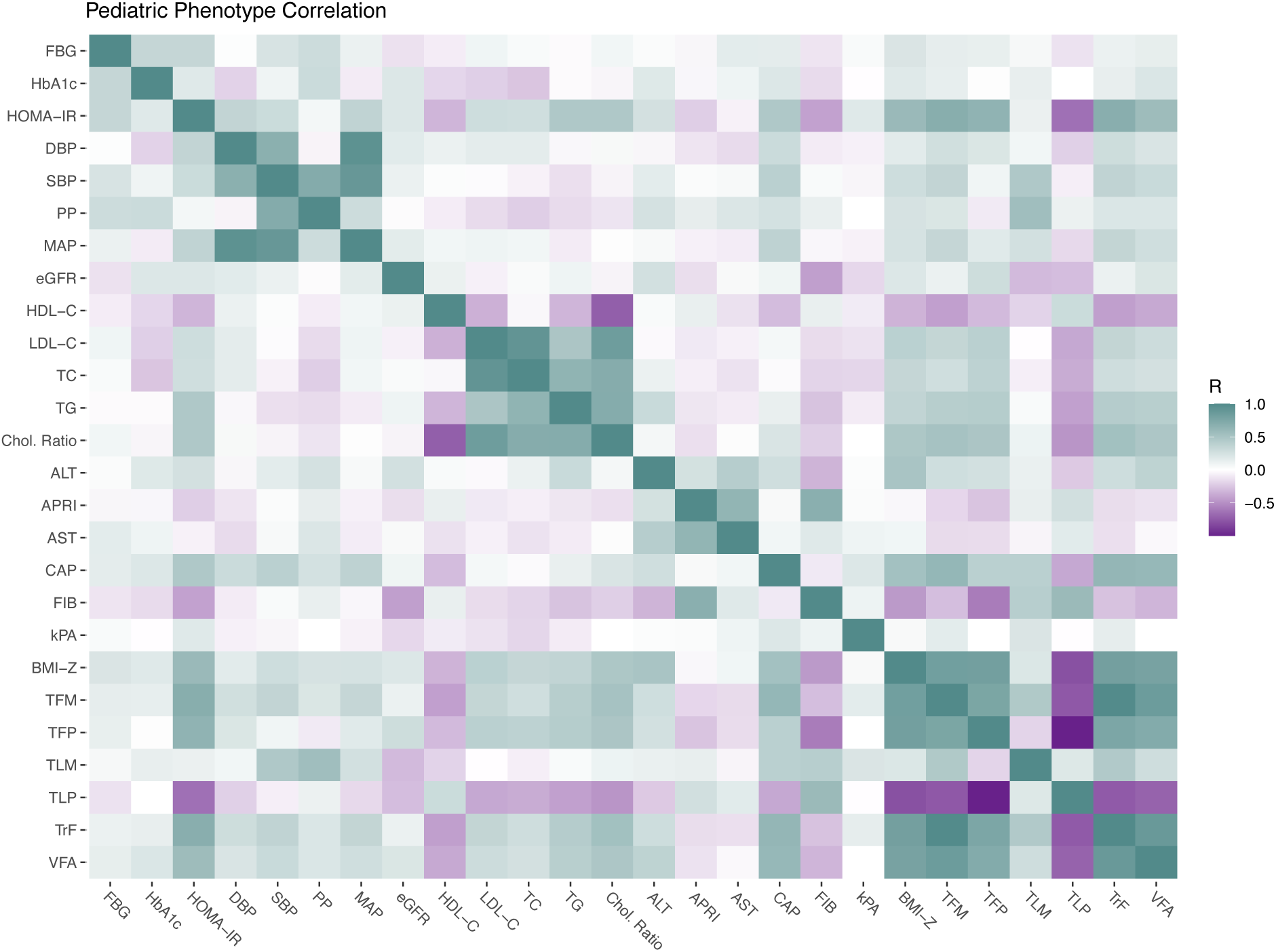
This heatmap shows the correlations between each pair of measured phenotypes in the pediatric study sample.

**Supplemental Figure 2.**
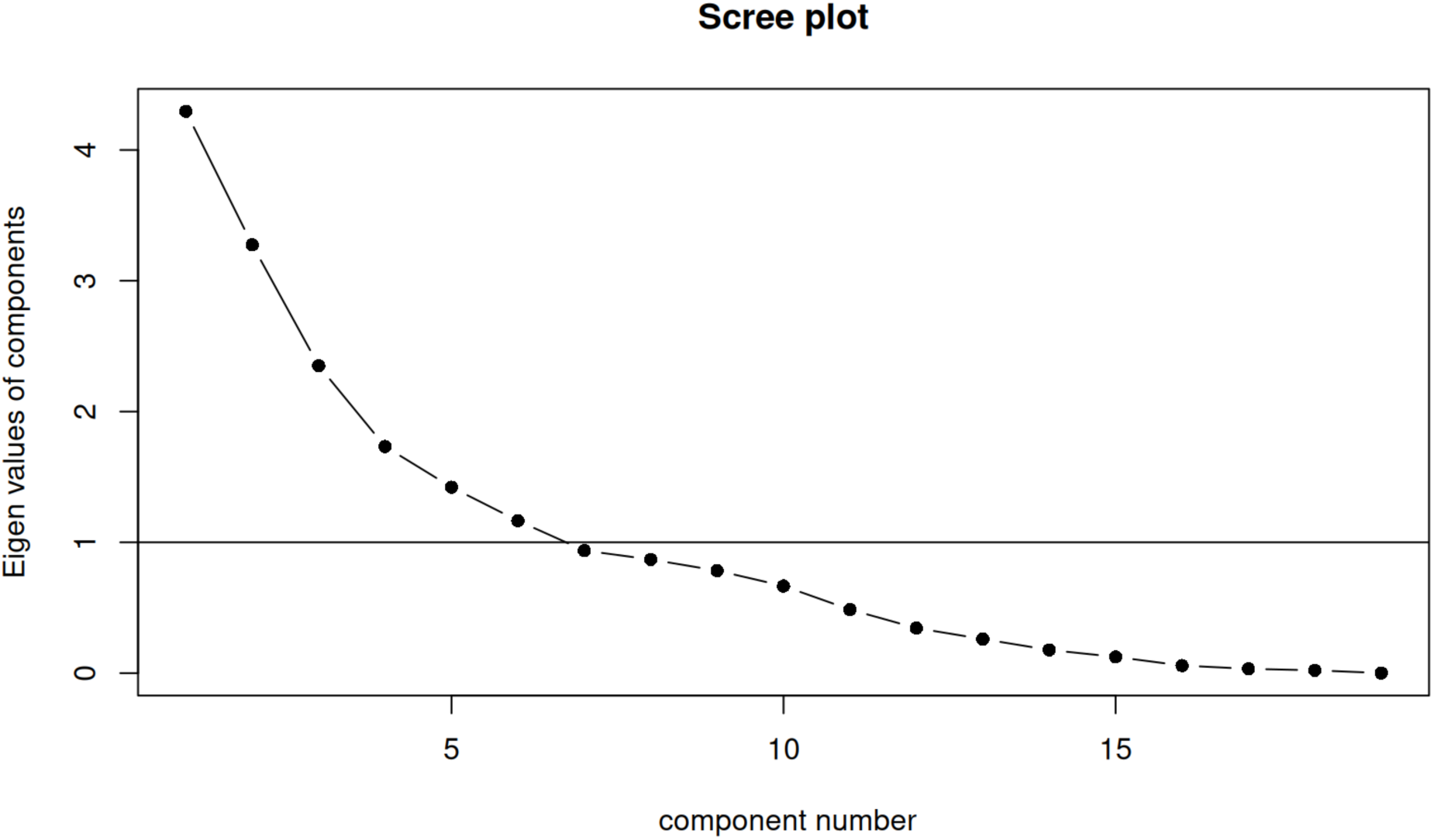
This scree plot indicates the eigenvalue of each component of the unrotated principal component analysis of the pediatric phenotype data. All components with eigenvalue > 1 (above the horizontal black line) were retained.

**Supplemental Figure 3.**
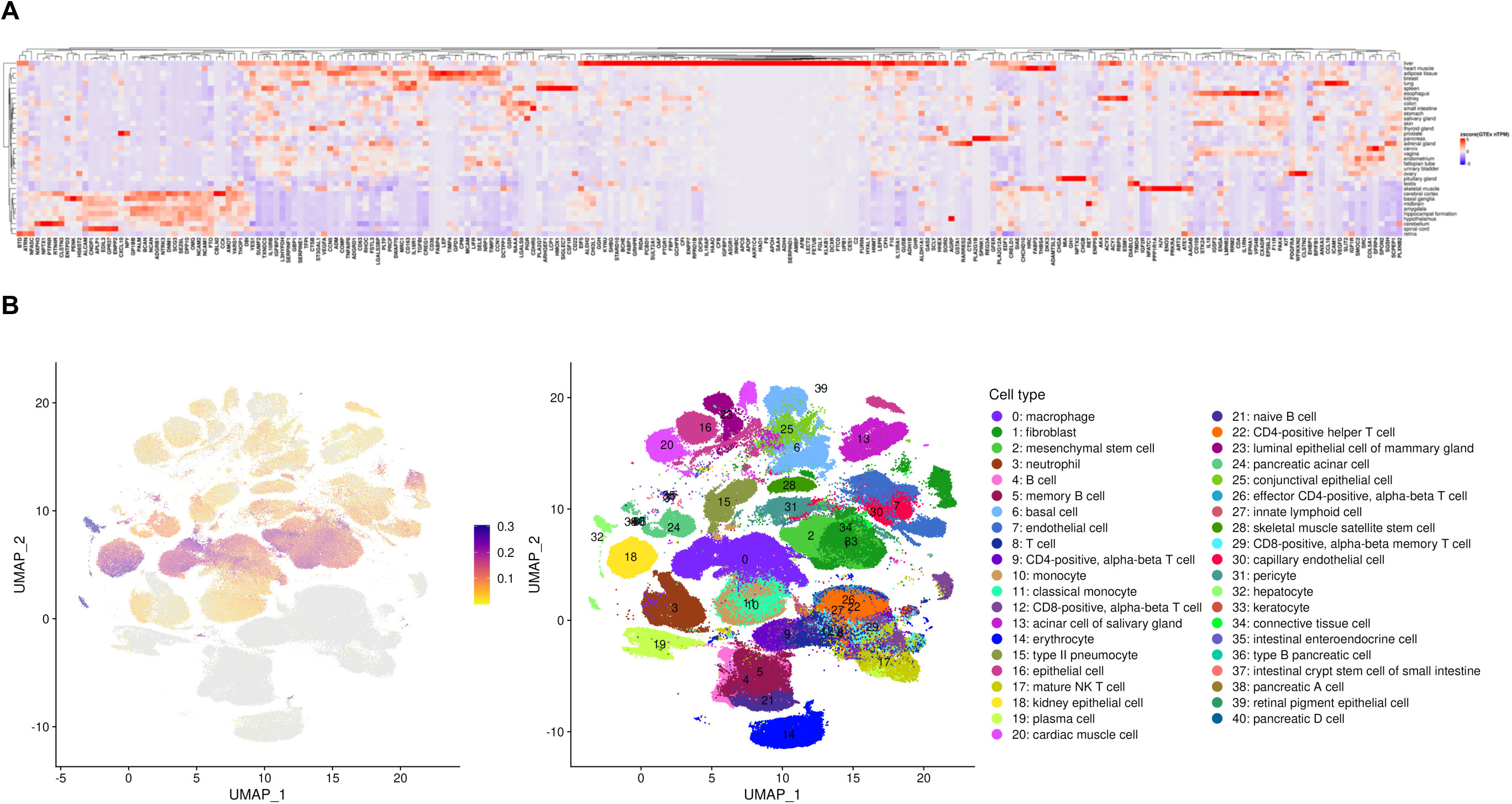
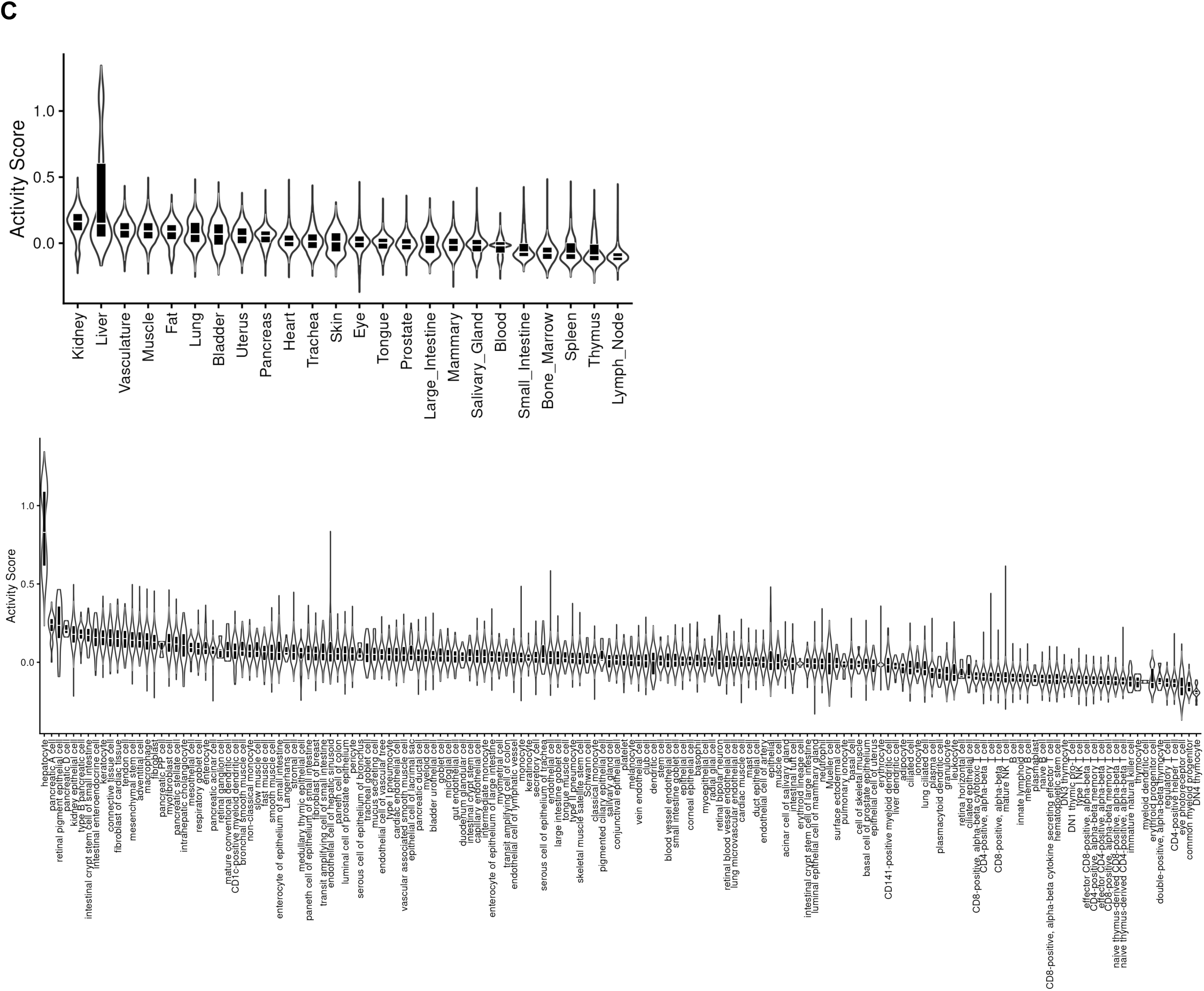
T**i**ssue **and cellular localization of the mutable pediatric CKMD proteome.** Here, the “mutable pediatric CKMD proteome” refers to proteins related to CKMD phenotypes in children that are also mutable with GLP-1RA therapy (both at 5% FDR, see **Methods**). Panel (A) displays tissue RNA-seq, demonstrating representation across liver, heart, adipose, kidney, and brain. Panel (B) displays a UMAP representation of *Tabula sapiens*, demonstrating transcriptional activity across different cell/tissue types. Panel (C) demonstrates tissue- and cell-specific transcriptional activity, highlighting similar tissues as (A) and specifically hepatocytes, pancreatic and renal cell types.

